# Understanding the effective reproduction number of *Plasmodium falciparum* malaria with seasonal variation at sub-national level in Nigeria

**DOI:** 10.1101/2024.04.29.24306577

**Authors:** Rabiu Musa, Abidemi Afeez, Olasupo Idowu Isaac, Mogbojuri Oluwaseun Akinlo, Samuel Abidemi Osikoya, Nwana Aaron Onyebuchi, Oniyelu Dolapo Oluwaseun, Olagbami Oluwafemi Samson, Bakare Emmanuel Afolabi

## Abstract

**Background:** With the highest burden in northern Nigeria, malaria is a vector-borne disease that causes serious illness. Nigeria contributed 27% (61.8 million) of malaria burden worldwide and 23% (94 million) of malaria deaths globally in 2019. Despite the fact that Nigeria has made a significant step in malaria elimination, the process has remained stagnant in recent years. The global technical strategy targets of reducing malaria death to less than 50 per 1000 population at risk was unachievable for the past 5 years. As part of the national malaria strategic plan of 2021-2025 to roll back malaria, it’s imperative to provide a framework that will aid in understanding the effective reproduction number (ℛ_*e*_) and the time dependent-contact rates *C*(*t*) of malaria in Nigeria which is quite missing in the literature.

**Methods:** The data of the reported malaria cases between January 2014 and December 2017 and demography of all the northern states are used to estimate *C*(*t*) and ℛ_*e*_ using Bayesian statistical inference. We formulated a compartmental model with seasonal-forcing term in order to account for seasonal variation of the malaria cases. In order to limit the infectiousness of the asymptomatic individuals, super-infection was also incorporated into the model.

**Results:** The posterior mean obtained shows that Adamawa state has the highest mean ℛ_*e*_ of 5.92 (95% CrI : 1.60-10.59) while Bauchi has the lowest 3.72 (95% CrI : 1.11-7.08). Niger state has the highest mean contact rate *C*(*t*) 0.40 (95% CrI : 0.08-0.77) and the lowest was Gombe 0.26 (95% CrI: 0.04-0.55 ). The results also confirm that there is a mosquito abundance and high reproduction number during the rainy season compared to the dry season. The results further show that over 60% of the reported cases are from the asymptomatic individuals.

**Conclusion:** This research continues to add to our understanding of the epidemiology of malaria in Nigeria. It is strongly advised that a complete grasp of the malaria reproduction number and the contact rate between human and mosquitoes are necessary in order to develop more effective prevention and control strategies. It will support the public health practitioner’s strategy and effective planning for malaria eradication.

## 1 Introduction

Malaria is a mosquito-borne infectious disease that affects humans and other vertebrates [1d, 2]. It is a life-threatening disease spread to humans by the infectious bite of female anopheles mosquitoes [2]. According to the World Health Organization (WHO), malaria caused 227 million cases and 409,000 deaths worldwide in 2019, with 94% (213 million) of those cases and 94% (386,000) of those deaths occurring in Africa [3].

In Nigeria, malaria is endemic and continues to be a serious public health concern [4]. Nigeria contributed 27% (61.8 million) of the worldwide burden of malaria and 23% (94 million) of all malaria deaths globally in 2019 [3, 5]. Nigeria is one of the six nations that contributed 51% of the world’s malaria cases in 2019 [3, 5]. The most susceptible population is the young children and pregnant women [6]. 30% of all admissions and 60% of all outpatient visits of those two categories are due to malaria [5, 6]. About 11% of maternal deaths and 30% of infant deaths in children under five are attributed to malaria in Nigeria [5]. The existing weak health system of the nation is further weakened by malaria, which also places a significant socioeconomic burden on the nation [7]. Malaria reduces the Gross Domestic Product (GDP) by 40% annually and costs the nation 480 billion Naira in direct and indirect medical expenses [7]. The rural north of Nigeria is particularly affected by malaria [8, 9]. In Borno, the number of cases of malaria reported in 2020 was 359,366 with 370 fatalities, whereas the number in 2019 was 330,587 with 240 fatalities [8]. According to routine statistics from DHIS2, there were 237,971 cases recorded in 2018 with 110 deaths, compared to 143 instances with 231 deaths in 2017. These numbers demonstrate that there was an increase in instances from 2017 to 2020. The prevalence of malaria has increased over time in other northern states of Nigeria [8]. The malaria prevalence rate among children under the age of five in Kaduna State, northern Nigeria, was 36.7%, which was higher than the national average of 27.7% [10]. In a news conference held in conjunction with World Malaria Day in 2022, the Kano state commissioner for health, Dr. Aminu Ibrahim Tsanyawa, said that 631 persons had died from malaria in the state in 2021. He claimed that in the first three months of 2022, 96 further fatalities were reported [11].

The symptoms of the illness, which include fever, headache, sweats, muscle pains, chills, exhaustion, nausea, vomiting, diarrhea, anemia, jaundice, and seizures, typically manifest in people 10–15 days after the infected mosquito has consumed its blood meal [12]. If an adequate treatment is not given promptly, uncomplicated malaria in patients with poor or no immunity may develop into severe malaria, which can be fatal. Malaria is curable, but if it is not effectively managed and treated, symptoms may return [2, 13].

Progress has halted in Nigeria in recent years despite increased funding for the universal scale-up of malaria control prevention, primarily through long-lasting insecticide-treated net (LLIN), seasonal malaria chemo-prevention (SMC), and treatment with artemisinin-based combination therapy (ACT) [14]. According to the World Health Organization’s Global Technical Strategy, there must be progress towards meeting the regional and global goals for 2020 and 2025 [14, 2]. One of the reasons is that the National Malaria Elimination Programs (NMEPs) from high malaria burden nations need to allocate their limited resources more wisely.

It is necessary for those states to customize the malaria interventions based on an adequate selection of intervention mixtures for particular risk areas in order to maximize success in those states, achieve malaria control, and proceed towards its elimination. This method calls for the sub-national stratification of a number of indicators of malaria risk derived from data on human behavior, parasite information, vector biology, and routine health surveillance. These indicators could be combined to produce an overall malaria risk score per distinct risk areas related to the local context in the country. To predict the impact that various tactics might have, mathematical modeling technique is very essential.

Nearly all viral, bacterial, and parasitic infections are known to have asymptomatic infectious phases, and new study is helping us understand how diseases spread from people who aren’t ill clinically [15]. Individuals without symptoms are less likely to seek treatment or adopt preventative measures [16, 17], while asymptomatic infected persons are frequently excluded from case definitions and control tactics in disease control programs [18]. Incorporating the severity of the asymptomatic class into the model used in fitting the malaria reported cases will be of great interest to the research community. It will make it easy to visually quantify the effects this category of infectious class have on the severity of malaria especially in the northern Nigeria.

Several mathematical models have been deployed to model the spread of malaria across the globe [19, 20, 21, 22, 23, 24, 25] among others. For example, [19] formulated a malaria transmission model with variables based on the climate. Using climatic data based on Representative Concentration Pathways (RCP) scenarios, they examine the effects of climate change on malaria transmission. Through the calculation of seasonal reproduction number and vectorial capacity under RCP scenarios and a sensitivity analysis of the model parameters, they analyze the possible danger of malaria outbreaks. They concentrate on the north of South Korea, which is where *Plasmodium vivax* malaria is prevalent. According to their findings, the likelihood of severe malaria outbreaks in the region will increase due to climate change.

In order to simulate malaria morbidity and mortality across Nigeria’s 774 LGAs under four potential intervention strategies from 2020 to 2030, an agent-based model of *Plasmodium falciparum* transmission was utilized by [20]. The scenarios included the currently implemented plan (business as usual), the NMSP with a coverage level of 80% or above, and two plans that were prioritized based on the resources available to Nigeria. Using monthly rainfall, temperature suitability index, vector abundance, pre-2010 parasite prevalence, and pre-2010 vector control coverage, LGAs were grouped into 22 epidemiological archetypes. Each archetype’s seasonality was parameterized using routine incidence data. According to their findings, if a business-as-usual policy were to be implemented, malaria incidence was predicted to rise by 5% and 9% between 2025 and 2030 compared to 2020 while mortality were predicted to stay the same. The NMSP scenario with 80% or more coverage of standard treatments, intermittent preventive therapy for babies, and expansion of seasonal malaria chemo-prevention (SMC) to 404 LGAs, as opposed to 80 LGAs in 2019, had the best intervention impact.

Modelling the effect of bed-net coverage on malaria transmission in South Sudan was studied by [21]. For the purpose of capturing the impact of LLINs on malaria transmission, they expanded an existing SIR mathematical model. With the aid of Markov Chain Monte Carlo (MCMC) techniques and a Bayesian approach, realistic parameter values for this model were established using the available data on malaria. Then, in order to develop a sub-national projection of malaria, they investigate the parasite prevalence on a sustained roll-out of LLINs in three different scenarios. They also determine the model’s reproductive number and investigate its sensitivity to the effectiveness and coverage of LLINs. They estimated the reproduction number based on the findings of the numerical simulation, which confirmed a significant rise in incidence cases in the absence of community intervention.

To the best of our knowledge, no study has estimated the effective reproduction number of malaria using the 2014-2017 *Plasmodium falciparum* malaria reported data. And no research has incorporated a time-dependent contact between human and mosquito in a mathematical model. This study seeks to use a compartmental mathematical model (incorporating seasonal variation, super infection and time-dependent contact rate) and reported malaria cases data to study the spread of *Plasmodium falciparum* malaria, estimate the time-dependent contact rate and the effective reproduction number of *Plasmodium falciparum* across the 20 northern states in Nigeria. The Bayesian statistical framework shall be used to estimate the time-dependent contact rates as well as the effective reproduction number for all the northern states.

The remaining sections are as follows: Section 2 contains the model structure, contact break, seasonality function, super-infection and description of the Bayesian inference framework. The data description and study area are discussed in section 3. The effective reproduction number was derived in section 4 while the discussion of results appears in section 5 followed by conclusion.

## 2 Methods

This section contains the foundation upon which this research is built. It entails an extensive discussion on the mathematical model and the population stratification.

### 2.1 Model Structure

In this study, a simple compartmental mathematical model is used to study the spread of malaria in northern Nigeria. The total human population denoted by 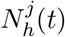 at state *j* and time *t* is subdivided into five com-partments of susceptible individuals 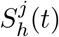, exposed individuals 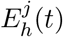, asymptomatic infectious individuals 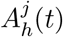, symptomatic infectious individuals 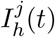 and lastly, the recovered individuals is denoted by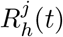.

Individuals in the exposed compartment do not transmit the disease [26, 27], while infectious individuals in 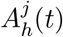 and 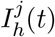 can transmit infection to their contacts at some rate. The recovery of the recovered individuals is assumed to be pseudo, ie. the recovery conferred on them is temporal because of the possibility of parasitaemia ruminants in their blood stream which makes them susceptible immediately after recovery. The total human population is denoted by

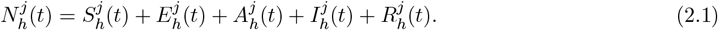

The schematic diagram of the model is given in Figure 1

**Figure 1:**
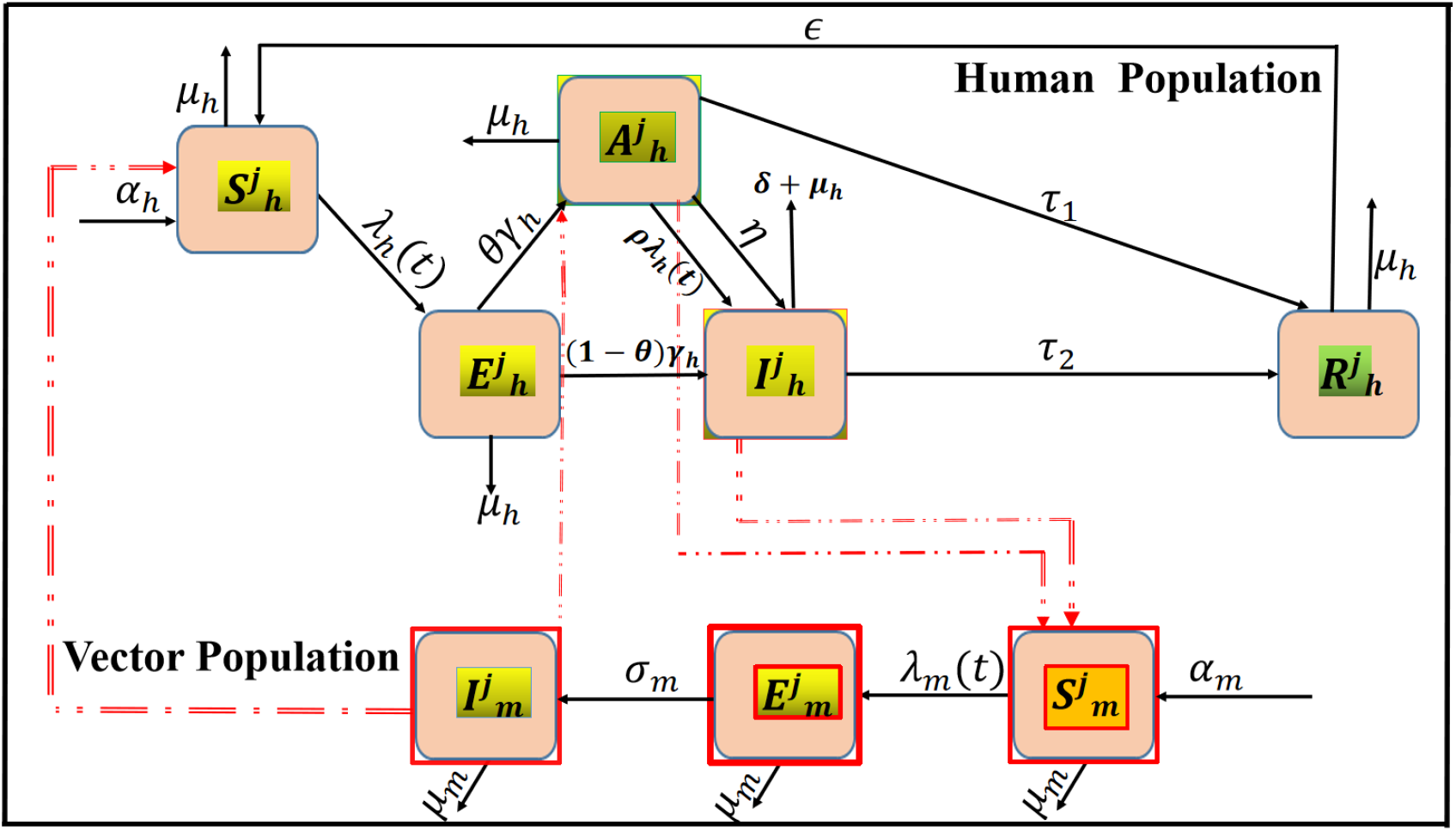
Schematic diagram of the model equation (2.3)-(2.6). The top part represents the human population in state j while the bottom part represents the vector population in state j. The red line denotes transfer of infection between both populations. All variables and parameters are as defined in Table 1.

**Figure 2:**
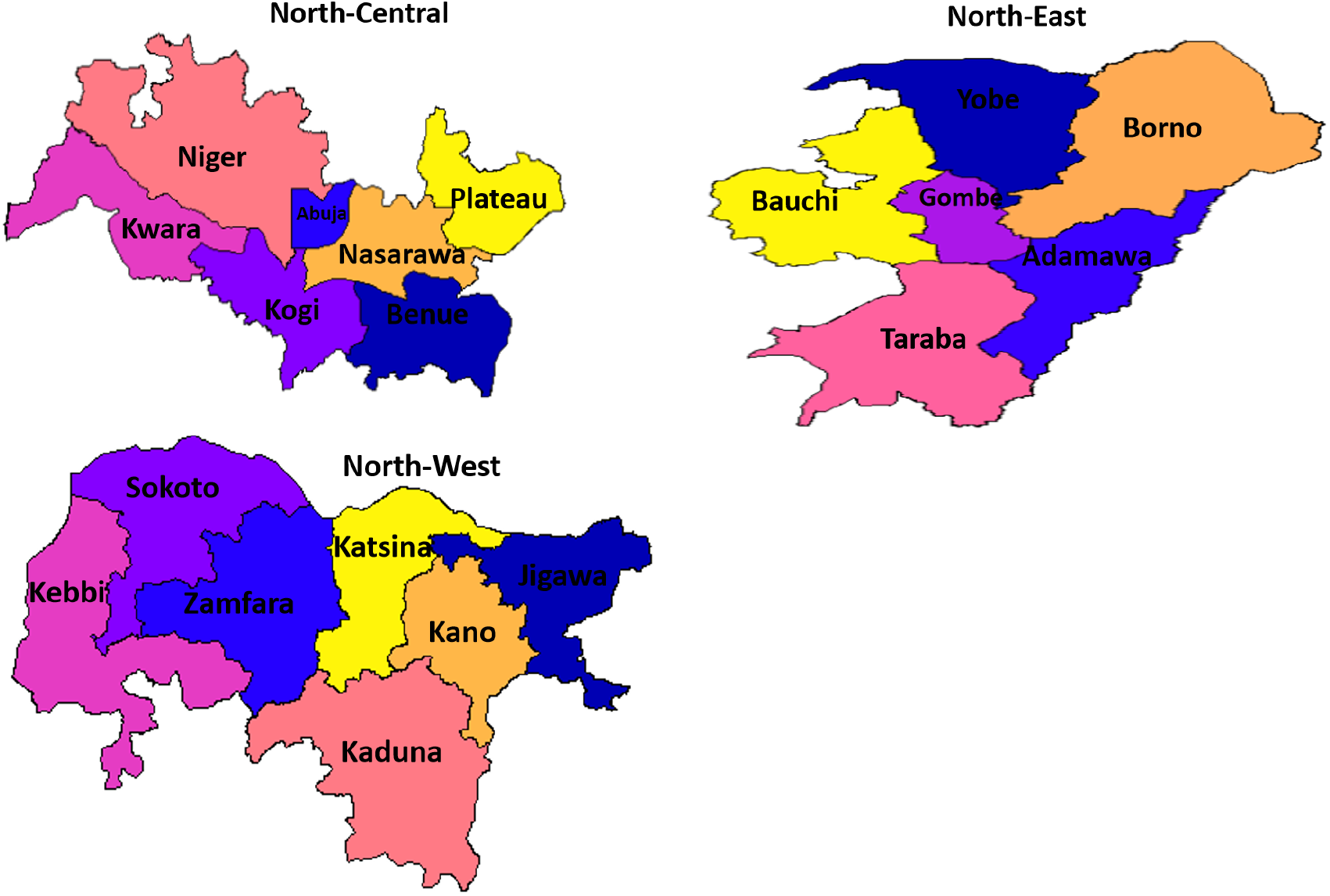
Map of the North-Central, North-Western, and North-Eastern Region of Nigeria. The North Central Region consists of Abuja, Benue, Kogi, Kwara, Nasarawa, Plateau and Niger states. The North Western Region consists of Jigawa, Kaduna, Kano, Katsina, Kebbi, Zamfara and Sokoto states. The North-Eastern Region Consists of Taraba, Borno, Gombe, Adamawa, Bauchi and Yobe states. The maps are generated using Shape-files and Geo-spatial data coded in R software.

**Figure 3:**
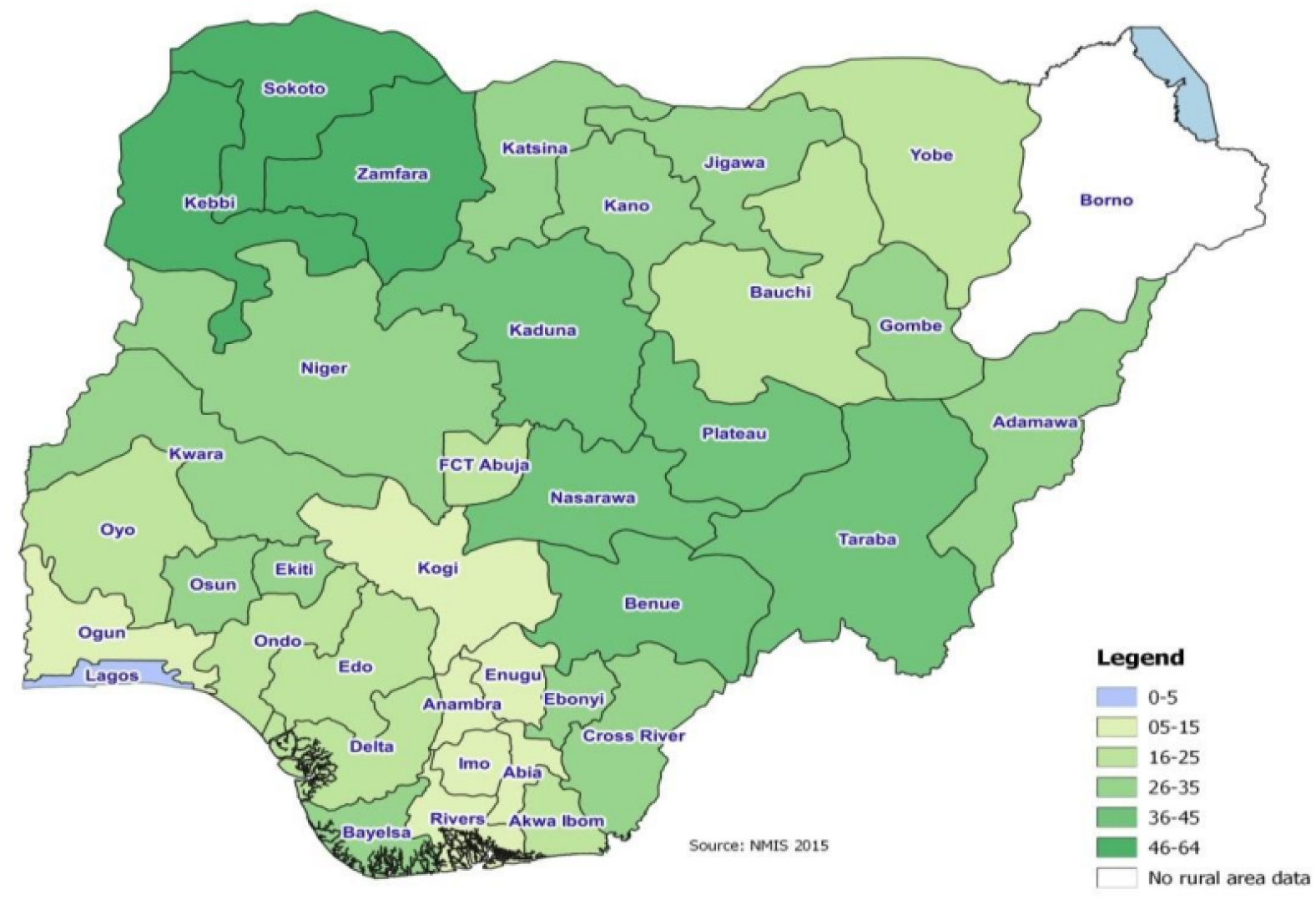
Map of Nigeria Showing the Spread of Malaria in Nigeria.[40]

**Figure 4:**
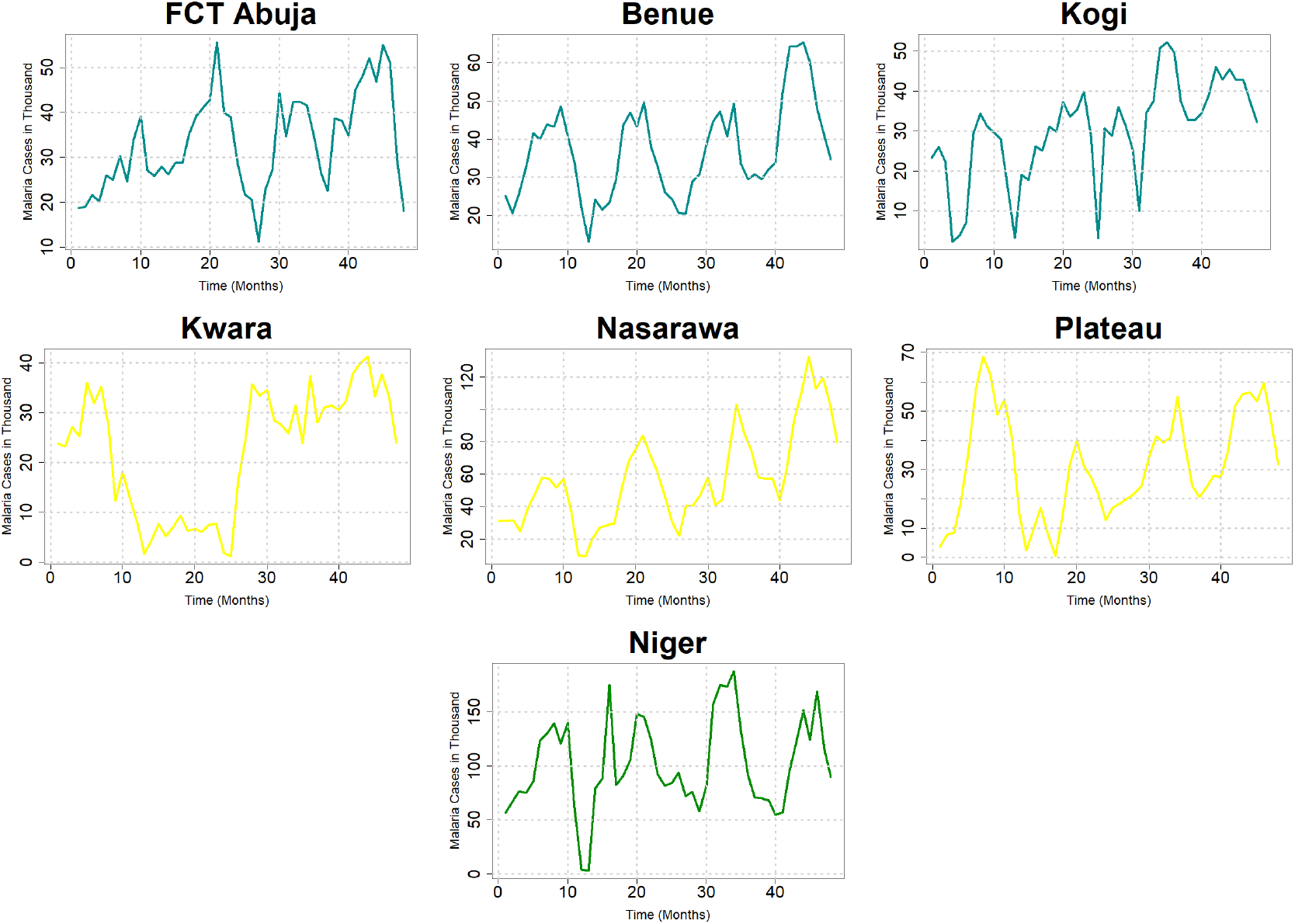
The times series plots of the monthly reported malaria cases for North Central Region consisting of Abuja, Benue, Kogi, Kwara, Nasarawa, Plateau and Niger states for the period of 4 years (January, 2014-December, 2017).

**Figure 5:**
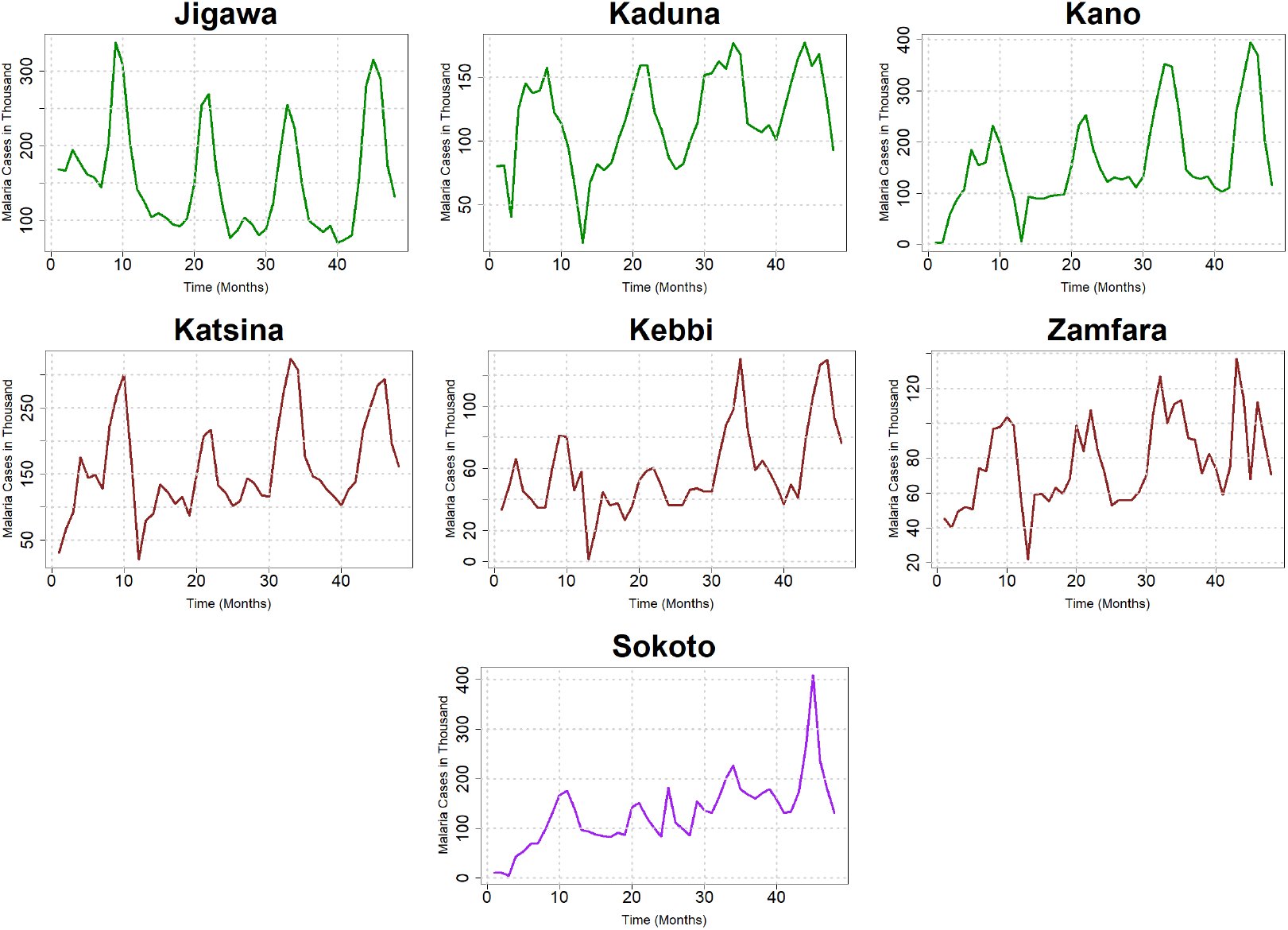
The times series plots of the monthly reported malaria cases for the North Western Region consisting of Jigawa, Kaduna, Kano, Katsina, Kebbi, Zamfara and Sokoto states for the period of 4 years (January, 2014-December, 2017).

The total mosquito population denoted by 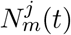 at state *j* and time *t* is subdivided into three compartments of susceptible mosquitoes 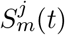, exposed mosquitoes 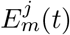 and infectious mosquitoes 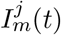. We do not consider the recovered compartment for the mosquito because of their short life cycle.

The total mosquito population is denoted by

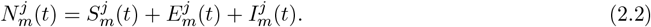

Majority of the malaria models in the literature assume that the contact rates between an infected mosquito and a susceptible human is always constant. Biologically, this assumption is not always true for several reasons. The number of contact received by a susceptible human varies depending on the temperature, relative humidity, season, location and time. Another reason the assumption is not always true is that malaria incidence in Nigeria is periodic following a specific seasonal pattern as a result of changes in the contact, biting and the entomological inoculation rates (EIR). On this backdrop, we incorporate a time-dependent contact rate and a seasonal pattern into the model in order to capture the trend of malaria in Nigeria.

**Table 1:** Model variables, parameters, descriptions and values.

| Variable | Description |  |  |
| --- | --- | --- | --- |
| $S_h^j$ | Susceptible human population in state $j$ | | |
| $E_h^j$ | Exposed human population in state $j$ | | |
| $I_h^j$ | Infectious human population in state $j$ | | |
| $R_h^j$ | Recovered human population in state $j$ | | |
| $S_m^j$ | Susceptible mosquito population in state $j$ | | |
| $E_m^j$ | Exposed mosquito population in state $j$ | | |
| $I_m^j$ | Infectious mosquito population in state $j$ | | |
| Parameter | Description | Value | Reference |
| $\gamma_h$ | Rate of transitioning from $E_h^j$ to $A_h^j$ ($\frac{1}{\gamma_h}$ is the incubation period) | $\frac{12}{30}$ month<sup>-1</sup> | [13] |
| $\tau_1, \tau_2$ | Recovery rates of both asymptomatic and symptomatic infectious classes respectively | $\frac{1}{24}, \frac{1}{9.5}$ month<sup>-1</sup> | [31] |
| $p$ | The probability of transmission of infection from an infectious human to a susceptible mosquito given that a contact occurred | 0.92<br>(Dimensionless) | Assumed |
| $\epsilon$ | The per capita rate of loss of immunity for some recovered humans | $1/2$ month<sup>-1</sup> | [26] |
| $\delta$ | The per capita disease induced death rate of humans | $0.00274$ month<sup>-1</sup> | |
| $\alpha_h$ | The per capita recruitment rate of humans | $N_h \mu_h$ | Derived |
| $\alpha_m$ | The per capita birth rate of mosquitoes | $N_m \mu_m$ | Derived |
| $\mu_m$ | The per capita death rate of mosquitoes | $1.87$ month<sup>-1</sup> | [32] |
| $\mu_h$ | The per capita death rate of humans | $\frac{1}{65 \times 12}$ month<sup>-1</sup> | [33] |
| $\sigma_m$ | Rate of transitioning from $E_m^j$ to $I_m$ ($1/\sigma_m$ is the incubation period) | $\frac{15}{7}$ month<sup>-1</sup> | [31] |
| $w$ | Period | $\frac{2\pi}{12}$ (Dimensionless) | Derived from data |
| $\phi_1$ | Amplitude | $0.8$ (Dimensionless) | Derived from data |
| $\phi_2$ | Vertical shift | $0.8$ (Dimensionless) | Derived from data |
| $s$ | Horizontal shift | $3.2$ (Dimensionless) | Derived from data |
| $\eta$ | Rate of transitioning from $A_h^j$ to $I_h^j$ | $\frac{3}{30}$ month<sup>-1</sup> | Assumed |
| $\kappa$ | The modification parameter for assumed reduction in transmission | $0.9$ (Dimensionless) | Assumed |
| $C(t)$ | Contact rate | $[0.26-0.40]$ | Estimated |
| $\theta$ | Proportion of exposed individuals that becomes asymptomatic | $0.60$<br>(Dimensionless) | Derived from data |
| $\rho$ | Super-infection rate | $0.0001$ month<sup>-1</sup> | Assumed |
| $\zeta_1, \zeta_2$ | Control measures for both human and vector populations respectively | $0.015, 0.02$<br>(Dimensionless) | Assumed |

#### Contact Break

The act of transforming a constant contact rate to a time-dependent type is referred to as contact break. According to Figure 6, it can be observed that the contact rate between mosquitoes and human varies with time which consequently leads to variations in the number of reported cases. It also worth noting that the number of reported cases is heavily dependent on the contact rates, number of bites and the probability of infection. For this reason, we considered a time-dependent contact rate *C*(*t*) in the model equation in order to account for such variation. The data was reported monthly between 2014 and 2017 which is equivalent to 48 months. We then assumed that there is changes in the contact rate bimonthly that is, every two months. This makes the contact rate changes 24 times between January 2014 and December 2017, hence we have *C*(*t*), 1 ≤ *t* ≤ 24. This approach has been used by many authors such as [28, 29].

**Figure 6:**
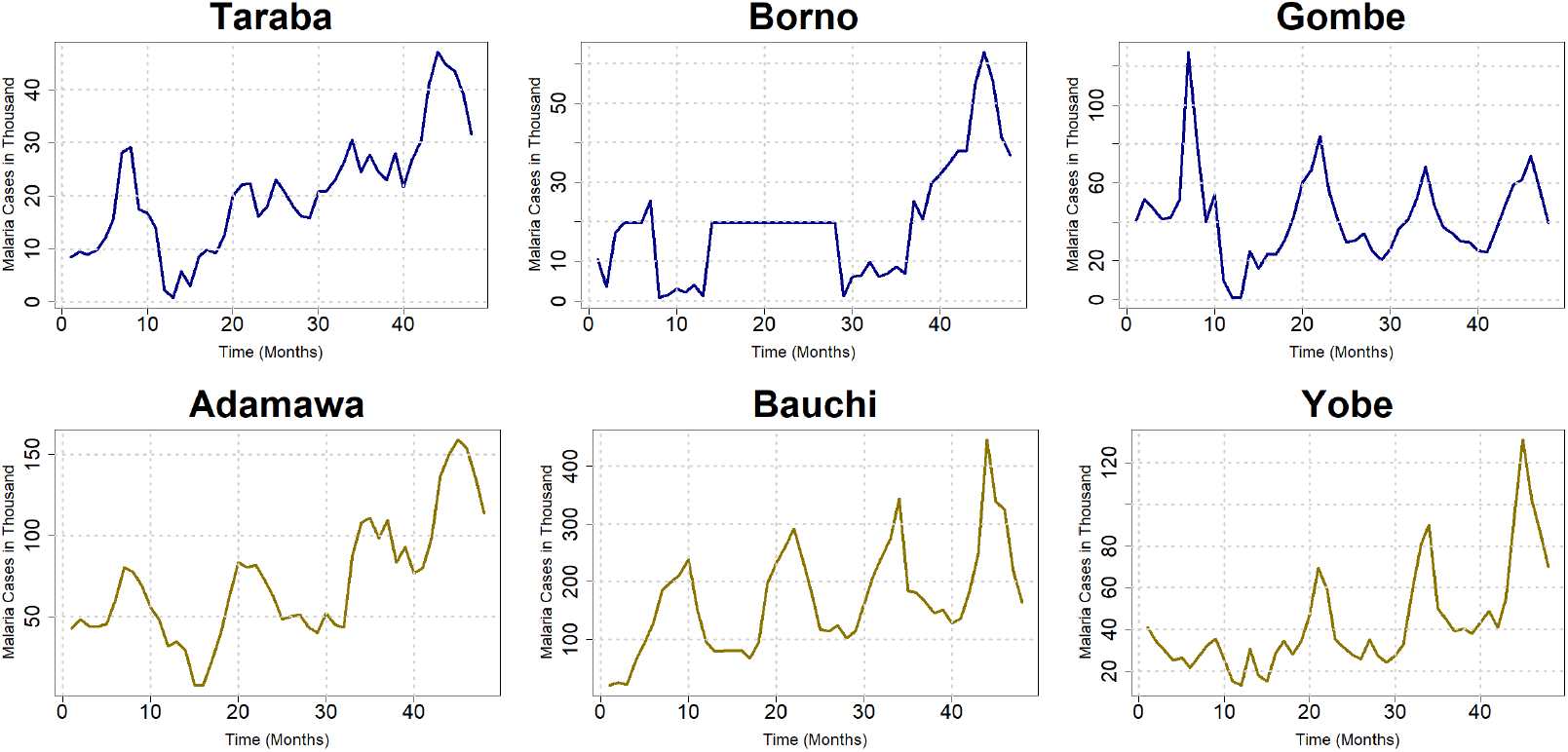
The times series plots of the monthly reported malaria cases for the North Eastern Region consisting of Taraba, Borno, Gombe, Adamawa, Bauchi, and Yobe states for the period of 4 years (January, 2014-December, 2017).

#### Seasonal Function

Nigeria has a year-round endemic malaria problem. Across the incredibly diverse ecological zones, there are significant differences in the transmission’s seasonality and intensity. In the savanna ecological zones of Derived Savannah, Guinea Savannah, Sudan Savannah, and Sahel Savannah, highly seasonal malaria occurs during and shortly after the time of strong rainfall (three to four months) [30]. According to the time series plots 6, it can be observed that the spread of malaria in Nigeria is seasonal. The peak is between August-November yearly and the lowest cases always recorded between December-April. In order to account for this seasonality, we incorporate a seasonal forcing term described in equation (2.4). The parameter *ϕ*_1_ which represents the amplitude accounts for the peak of the reported malaria cases. The trigonometric functions account for the rise and fall of the malaria cases. After the fitting was done, we discovered that the fitted curve was unable to capture the initial spread after take-off of the malaria cases, hence the need to introduce the horizontal phase shift *s*. We shifted the fitted curve by distance *s* = 0.8 and vertical phase shift, *ϕ*_2_ = 0.8 in order to avert this problem.

#### Human Population

We incorporate birth and death rates in the model to capture demography of the northern states. The susceptible human population is recruited either by birth or migration at a rate *α*_*h*_. It is assumed biologically that the susceptible human population gets infected by an infectious female anopheles mosquito at a rate *λ*_*h*_ after a successful bite and transit to the exposed compartment. After the waning of the natural or treatment-induced immunity, the fully recovered humans become susceptible to the new infection at a rate *ϵ* while natural death occurs at a per capita death rate *µ*_*h*_. After the successful transfer of infection from the infectious female anopheles mosquito, the exposed human becomes infectious and transit to the asymptomatic infectious compartment at the rate *θγ*_*h*_ while the remaining exposed individuals transit to the symptomatic class at the rate (1 − *θ*)*γ*_*h*_. The rate of transitioning between the asymptomatic and symptomatic class is given by *η*. The disease-induced death occurs at the rate *δ* while the infectious individuals *A*_*h*_, *I*_*h*_ recover at the rate *τ*_1_, *τ*_2_ respectively. The set of ordinary differential equation governing the human population is given by

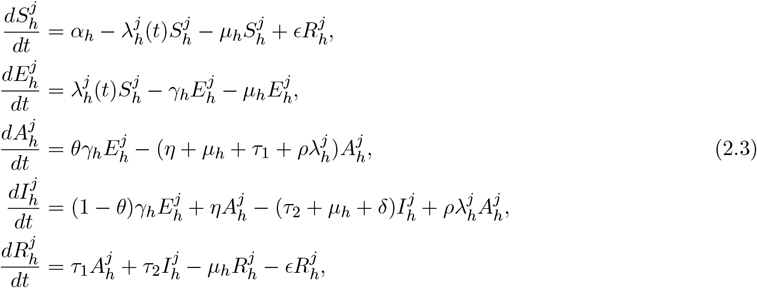

where all state variables and model parameters are assumed to be non-negative.

The force of infection is also given by

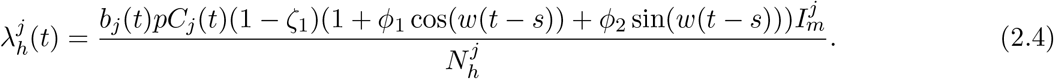

The force of infection contains several components. The time-dependent contact rates between the infectious human and susceptible mosquito (and vice versa ) is denoted by *C*_*j*_(*t*) where 1 ≤ *t* ≤ 24 for state *j*, the total number of bites is denoted by *b*_*j*_ for state *j* while the probability of transmission is given by *p*. The remaining part of the expression is the seasonal-forcing term where *ϕ*_1_ is the amplitude of the periodic graph, *s* is the horizontal phase shift, *ϕ*_2_ is the vertical phase shift, 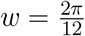 is the period of oscillation and 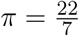.

#### Super-Infection

In order to limit the asymptomatic infectious period, we incorporated the dynamics of super-infection (denoted by 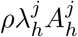) on the asymptomatic class so that they can become symptomatic quickly. A cell that has already been infected by one virus can become super-infected when it is later infected by a different strain of that virus or another virus. Reduction of the asymptomatic-infectious period can go a long way in eliminating or possibly eradicating malaria. Since variety of control measures have been in place in the northern Nigeria during and before the reported malaria cases data between January 2014 and December 2017, we include parameters *ζ*_1_, *ζ*_2_ denoting control measures implemented in the northern states for both human and vector population respectively.

### Mosquito Population

The susceptible mosquito population is generated through birth at the rate *α*_*m*_, they become infected at the rate *λ*_*m*_ (generally considered as the force of infection) while natural death occurs at the rate *µ*_*m*_. The exposed mosquitoes become infectious at the rate *σ*_*m*_. The total mosquito population is given by the equations

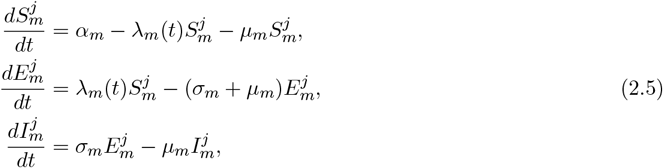

where all state variables and model parameters are assumed to be non-negative.

The force of infection is also given by

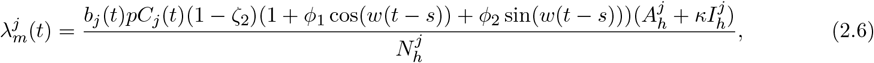

where *κ* denotes the assumed reduction in the transmission rate of the symptomatic individuals.

Table 1 contains the parameter values, model variables and their description for both populations. Majority of the parameter values are obtained from the literature, while the time-dependent contact rate *C*(*t*) is estimated for each state from the monthly reported malaria cases using Bayesian inference. The effective reproduction number for each state is then computed using (4.2) and the estimated time-dependent contact rate. The estimated time-dependent contact rates, initial condition and the effective reproduction number for all the northern states as well as their credible intervals are available on Table 3.

### 2.2 Bayesian Inference

The malaria model (2.3) - (2.6) with periodic function (2.4) capable of capturing the seasonal variation in the spread of malaria and time-dependent contact rates as described in the aforementioned equations was fitted to the monthly reported malaria cases in the northern Nigeria spanning between 2014 and 2017. The data set was collected from the national malaria elimination program (NMEP) [34] and can be accessed upon request since its not publicly available. The Bayesian inference framework and the Rstan package [35] in R version 4.3.1 [36] was used to obtain the posterior mean. This probabilistic framework allows us to take into account our prior knowledge when estimating the model parameters and also allows us to evaluate probabilistic statements about the data in light of the model. The sampling distribution also known as likelihood involves the data generating process given the model parameters. The posterior distribution involves the process of estimating plausible parameter values given the data. The fit summary contains useful information such as the mean, sample size, standard deviation, credible interval, the convergence criterion Rhat etc. In general, the Bayesian frame-work is built upon the relationship

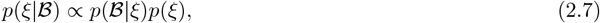

where *p*(*ξ*|ℬ) is the likelihood, *p*(ℬ|*ξ*) is the posterior distribution, and *p*(*ξ*) is the prior. For each northern states at sub-national level in Nigeria, the likelihood is constructed as

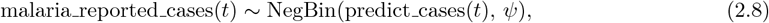

where malaria_reported_cases is the monthly reported cases of malaria in the northern Nigeria at time *t* ( measured in months), predict cases(*t*) is the predicted monthly malaria cases computed from the model (2.3)-(2.6), NegBin(·) is the negative binomial distribution function in RStan, and *ψ* is the over-dispersion parameter.

To ascertain the fact that our model (2.3) - (2.6) is correctly coded in the Stan function and to ensure that the proposed model is properly validated, synthesized cases data was generated using the proposed model with known parameter values. The model’s ability to estimate these parameter values after fitting the model to the synthesized data was examined. The obtained posterior distributions were inspected for biases and coverage of the true parameter values used to generate the synthesized data (see Figures S1 and S2 in the appendix). The adaptive Hamiltonian Monte Carlo method No-U-Turn sampling (NUTS) in RStan with 1,000 iterations and 4 chains was used to conduct all analyses and estimations reported in this research. Details of these methods can be found in [37, 38]

## 3 Materials

### 3.1 Data

The data used for this research is the malaria incidence data obtained from the National Malaria Elimination Programme (NMEP) [34] between January 2014 and December 2017 which is equivalent to 48 months. The data contains 43 variables including information on the anti-natal attendance, age-distribution of malaria cases, long-lasting insecticide-treated net (LLIN), immunized children, Malaria RDT tested positive, Malaria microscopy tested positive, Pregnant women with clinical malaria among others. We selected those states based on the availability of data and mostly because the northern Nigeria is the most affected region in Nigeria. In Figure 6, the 10th month corresponds to October 2014, the 20th month corresponds to August 2015, 30th month corresponds to June 2016, 40th month corresponds to April 2017 and so on.

In order to reduce the noise of the data and most importantly, to ease the analysis, we scaled down the number of reported cases in thousand and plotted them against time as shown in the Figures. There is no missing data for the period of time considered. In our model (2.3)-(2.6), the number of predicted monthly cases of malaria is computed in Stan as the number of asymptomatic individuals 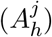 transitioning to the infectious compartment 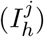.

We also used the demography of the northern states as the initial condition of the total human population. The population size of each northern states is based on the 2006 national population Census of Nigeria as well as their 2019 projected populations published by the National Bureau of Statistics [33] in the year 2020.

### 3.2 Study Area and Malaria Spread

A major public health risk in Nigeria is malaria, which is predicted to cause 68 million cases and 194 000 deaths worldwide by 2021 [9]. With roughly 27% of the world’s malaria cases, Nigeria has the largest global malaria burden. Year-round, there is a risk of transmission anywhere in the nation. However, the northern region of the nation accounts for the greatest rates of malaria [9, 39]. Our study area in this research is the northern states in the northern Nigeria which includes Bauchi, Borno, Kaduna, Kano, Kogi, Sokoto, Zamfara etc as appeared on Table 2. The map of the selected states are shown in Figure 2 below.

**Table 2:** List of the northern states that constitutes our study area and their population sizes. The population size is based on the 2006 national population Census of Nigeria as well as their 2019 projected populations, which were published by the National Bureau of Statistics [33].

| Northern State | Population<br>size | Region |
| --- | --- | --- |
| Abuja | 2,702,443 | **North-Central** |
| Benue | 5,787,706 |  |
| Kogi | 4,153,734 |  |
| Kwara | 3,259,613 |  |
| Nasarawa | 2,632,239 |  |
| Plateau | 4,400,974 |  |
| Niger | 6,220,617 |  |
| Jigawa | 6,779,080 | **North-West** |
| Kaduna | 8,324,285 |  |
| Kano | 14,253,549 |  |
| Katsina | 9,300,382 |  |
| Kebbi | 5,001,610 |  |
| Zamfara | 5,317,793 |  |
| Sokoto | 5,863,187 |  |
| Taraba | 3,331,885 | **North-East** |
| Borno | 5,751,590 |  |
| Gombe | 3,623,462 |  |
| Adamawa | 4,536,948 |  |
| Bauchi | 7,540,663 |  |
| Yobe | 3,398,177 |  |

**Table 3:** Estimated time-dependent contact rates, initial condition and the effective reproduction number for all the northern states as well as their credible intervals.

| Northern States | Mean<br>estimated<br>contact rate | Estimated<br>Initial<br>Condition $I_h^j(0)$ | Mean Estimated $\mathcal{R}_e$ | Region |
| --- | --- | --- | --- | --- |
| Abuja | 0.30 (95% CrI : 0.07- 0.55) | 266 (95% CrI :210-299) | 4.85 (95% CrI : 1.11-8.76) | North Central |
| Benue | 0.27 (95% CrI : 0.07-0.48) | 286 (95% CrI :251-299) | 5.25 (95% CrI : 1.42-9.38) |  |
| Kogi | 0.27 (95% CrI : 0.02-0.55) | 264 (95% CrI :175-287) | 5.32 (95%CrI 0.45-10.98) |  |
| Kwara | 0.33 (95% CrI : 0.09-0.58) | 278 (95% CrI :237-298) | 4.88 (95% CrI : 1.36-8.38) |  |
| Nasarawa | 0.28 (95% CrI : 0.05-0.53) | 280 (95% CrI :238-299) | 5.39 (95% CrI : 0.97-10.28) |  |
| Plateau | 0.30 (95% CrI : 0.04-0.61) | 88 (95% CrI :19-189) | 4.50 (95% CrI : 0.67-9.17) |  |
| Niger | 0.40 (95% CrI : 0.08-0.77) | 286 (95% CrI :259-299) | 4.25 (95% CrI : 0.86-8.09) |  |
| Jigawa | 0.37 (95% CrI : 0.07-0.72) | 290 (95% CrI :272-299) | 4.77 (95% CrI : 0.96-9.35) | North-West |
| Kaduna | 0.31 (95% CrI : 0.06-0.61) | 288 (95% CrI :264-299) | 5.25 (95% CrI : 1.01-10.49) |  |
| Kano | 0.31 (95% CrI : 0.06-0.61) | 202 (95% CrI :99-290) | 4.70 (95% CrI: 0.99-9.08) |  |
| Katsina | 0.35 (95% CrI : 0.08-0.67) | 286 (95% CrI :247-299) | 5.03 (95% CrI : 1.14-9.66) |  |
| Kebbi | 0.33 (95% CrI: 0.06-0.64) | 285 (95% CrI :248-299) | 4.98 (95% CrI: 0.89-9.71) |  |
| Zamfara | 0.31 (95% CrI: 0.06-0.61) | 285 (95% CrI :254-298) | 4.63 (95% CrI: 1.01-8.48) |  |
| Sokoto | 0.31 (95% CrI: 0.06-0.57) | 175 (95% CrI :92-271) | 4.93 (95% CrI: 0.64-9.31) |  |
| Taraba | 0.31 (95% CrI: 0.04-0.59) | 285 (95% CrI :254-298) | 4.93 (95% CrI: 0.64-9.31) | North-East |
| Borno | 0.36 (95% CrI : 0.05-0.69) | 194 (95% CrI :71-294) | 4.71 (95% CrI : 0.62-8.96) |  |
| Gombe | 0.26 (95% CrI: 0.04-0.55 ) | 274 (95% CrI :205-299) | 5.37 (95% CrI : 0.74-11.31) |  |
| Adamawa | 0.27 (95% CrI: 0.07-0.48 ) | 286 (95% CrI :251-299) | 5.92 (95% CrI : 1.60-10.59) |  |
| Bauchi | 0.33 (95% CrI : 0.09-0.62) | 277 (95% CrI :237-291) | 3.72 (95% CrI : 1.11-7.08) |  |
| Yobe | 0.31 (95% CrI: 0.07-0.60) | 285 (95% CrI :254-298) | 4.84 (95% CrI: 1.10-9.33) |  |

### 3.3 Malaria Cases Time-Series Plot for the Study Area

The time series plots of the monthly reported malaria cases for all the northern states for the period of 4 years (January, 2014-December, 2017) are shown in Figure 6. In those plots, we note the following points:

1. It can be observed from all the plots that malaria has been endemic in Nigeria far before the start of the year 2014 simply because cases were reported in thousands on a monthly basis. In some states such as Bauchi, Abuja, Katsina, Kano and Sokoto, there was a sharp increase in the number of reported cases at the beginning of January 2014. This trend continues up to the start of November 2014 for Bauchi state which was immediately followed by a sharp decline in the number of reported cases. For Sokoto, Benue, Katsina, Kano etc, the initial sharp increase was followed by a short and sharp decline followed by some seasonal variations. For other states like Borno, Benue, and Zamfara, the initial reported cases declined sharply and quickly before experiencing a sharp increase in the number of reported cases. The trend in Kaduna, Nasarawa and Jigawa states maintained a uniform number of reported cases before experiencing a sharp decline (in Nasarawa and Kaduna) while that of kogi state had a sharp and short increase followed by a sharp decline in the number of reported cases.
2. The reported cases show a sharp decline towards the end of the 48th month ie. (towards the end of 2017) which makes it look like the outbreak of malaria in Nigeria is almost eradicated. It only declined because of the season. It picks up immediately at the beginning of the following season in 2018.
3. The reported cases of malaria exhibits some variational pattern like wave which shows that the incidence and spread of malaria in Nigeria do not follow a uniform or constant pattern. This can be attributed to variations in the number of bites, contact and transmission rates of malaria. This variational pattern or wave can be easily seen in Kogi (between the 12th and 20th month), Kaduna (between 30th and 35th month), Sokoto (within the first 10 months) among others.
4. The spread of malaria in Nigeria demonstrates strong seasonality. It rises and falls at some times in a year based on temperature, weather condition, humidity among others. Significant differences exist between locations and between years in the type and degree of seasonality. The most fundamental characteristics of malaria epidemiology are its regional spread and temporal fluctuation in transmission.

## 4 The Effective Reproduction Number

The disease-free equilibrium of (2.3)-(2.6) is given by

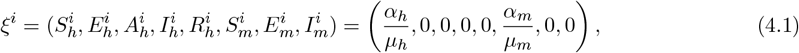

Where 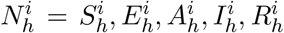 , and 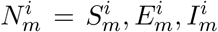. The reproduction number is computed using the next generation matrix operator [41, 42] on (2.3)-(2.6) as used by [43, 44]. The matrix for the rates of new infections ℱand that of transfer of infections, 𝒱 at the DFE are given by

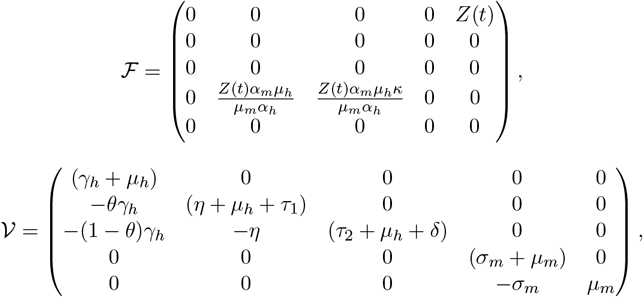

where *Z*(*t*) = (1 + *ϕ*_1_ cos(*w*(*t* − *s*)) + *ϕ*_2_ sin(*w*(*t* − *s*)))*b*_*j*_*pC*_*j*_(*t*).

The next generation matrix is defined by ℱ𝒱^*−*1^. Evaluating the spectral radius (dominant eigenvalue) of the next generation matrix, the reproduction number is obtained as

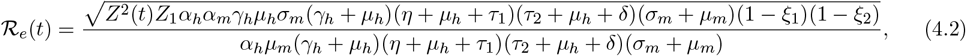

Where

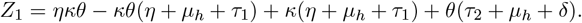

The estimated number of cases in a community where everyone is vulnerable to infection that can be directly attributed to one case is known as the basic reproduction number [43, 44, 45]. If some individuals in the population are immune or no more susceptible, then the basic reproduction number becomes the effective reproduction number ℛ_*e*_. ℛ_*e*_ is the center of all mathematical modeling work, and without it, it is impossible to estimate the spread or control of illnesses.

Since the effective reproduction number is periodic, it is very instructive to properly define the average of a periodic function over its period. Hence, we have the following definition.

**Definition 1**. *If the effective reproduction number ℛ*_*e*_(*t*) *is a periodic function of period ω, then the mean or average of ℛ*_*e*_ *over the period is given by*

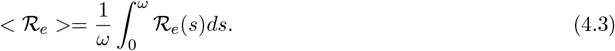

Now, we have the following preposition.

**Preposition 1**. *If ℛ*_*e*_ *is a periodic function of period ω, then*

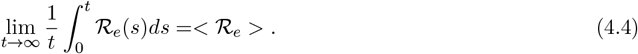

*Proof*. Let *t* ∈ [*nω*, (*n* + 1)*ω*]. Then *t* = *nω* + *ϵ*, where *ϵ* ∈ [0, *ω*]. Using the properties of definite integral in integral calculus, we have

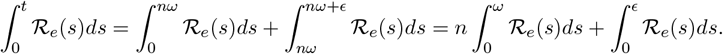

Dividing through by *t*, taking *t* = *nω* + *ϵ* on the right-hand side to have

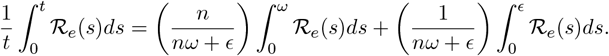

Taking the limit of both sides as *n* → ∞ to have

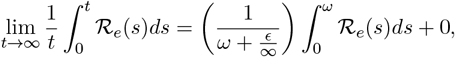

which gives Hence,

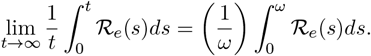

Hence,

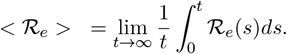

The interpretation of this result is that the time-dependent effective reproduction number ℛ_*e*_ over the period of consideration (January 2014-December2017) is the mean reproduction number for each state over the same period.

## 5 Results and discussion

In this section, we shall present the main findings of this research followed by discussion of results.

### 5.1 Model Calibration and Parameter Estimation

In this research, we adopted a Bayesian statistical framework to condition our inference about the reproduction number ℛ_*e*_ in (4.2), the time dependent contact rate *C*(*t*), the force of infection (2.4) and the predicted case counts on the number of reported cases. The predicted number of case counts was related to the reported malaria data through a negative binomial observation model with *ϕ* as its over-dispersion parameter. We implemented a weakly informative prior on *C*(*t*), ℛ_*e*_, and *λ*_*h*_ using a uniform distribution. A log-normal distribution was used in the estimation of the initial condition while an exponential distribution was used for the over-dispersion parameter.

The Bayesian inference framework and the Rstan package [35] in R version 4.3.1 [36] was used to obtain the posterior mean and the sampling distribution also known as likelihood. Stan implements the adaptive Hamiltonian Monte Carlo method No-U-Turn sampling (NUTS) algorithm for Bayesian statistical inference. We started the analysis sampling from 4 chains with 1000 iterations each discarding the first 500 as warmup. The diagnostic plots such as trace plots and density plots were used to examine chain convergence while ensuring that the 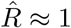 (Rhat is the potential scale reduction factor on split chains. At convergence, Rhat=1) . We also ensure that the effective sample size is greater or equal to 200. The uncertainties were presented using quantile-based credible intervals. The approach was validated using simulated data and Figures are presented in S1 and S2 in the appendix A .

### 5.2 Model Validation

The model validation involves the use of synthesized data extracted from the ODE solution of the model equations. This data is used to test the ability of the model to recover the posterior mean. The model showed a good fit (see Figures S1 and S2 in the appendix A ) and was able to recover *ϵ* = 0.5 that was used in the ODE solution. This shows that the inference, posterior mean and estimates of our proposed model are reliable.

The model fits for all the northern states are presented in Figures 7 -9. It can be seen that our model was able to capture the malaria spread in the northern Nigeria. The seasonality function was also able to match the rise and fall as well as the oscillation in the reported cases. There was an exception in the fit of Jigawa, Gombe, Niger, Zamfara, Yobe and Adamawa states due to a slight mismatch in the fit of those states during the initial disease spread. One possible reason for this is that the quality of the data in those states maybe poor. There maybe over-reporting of the true cases in those states during the initial spread.

**Figure 7:**
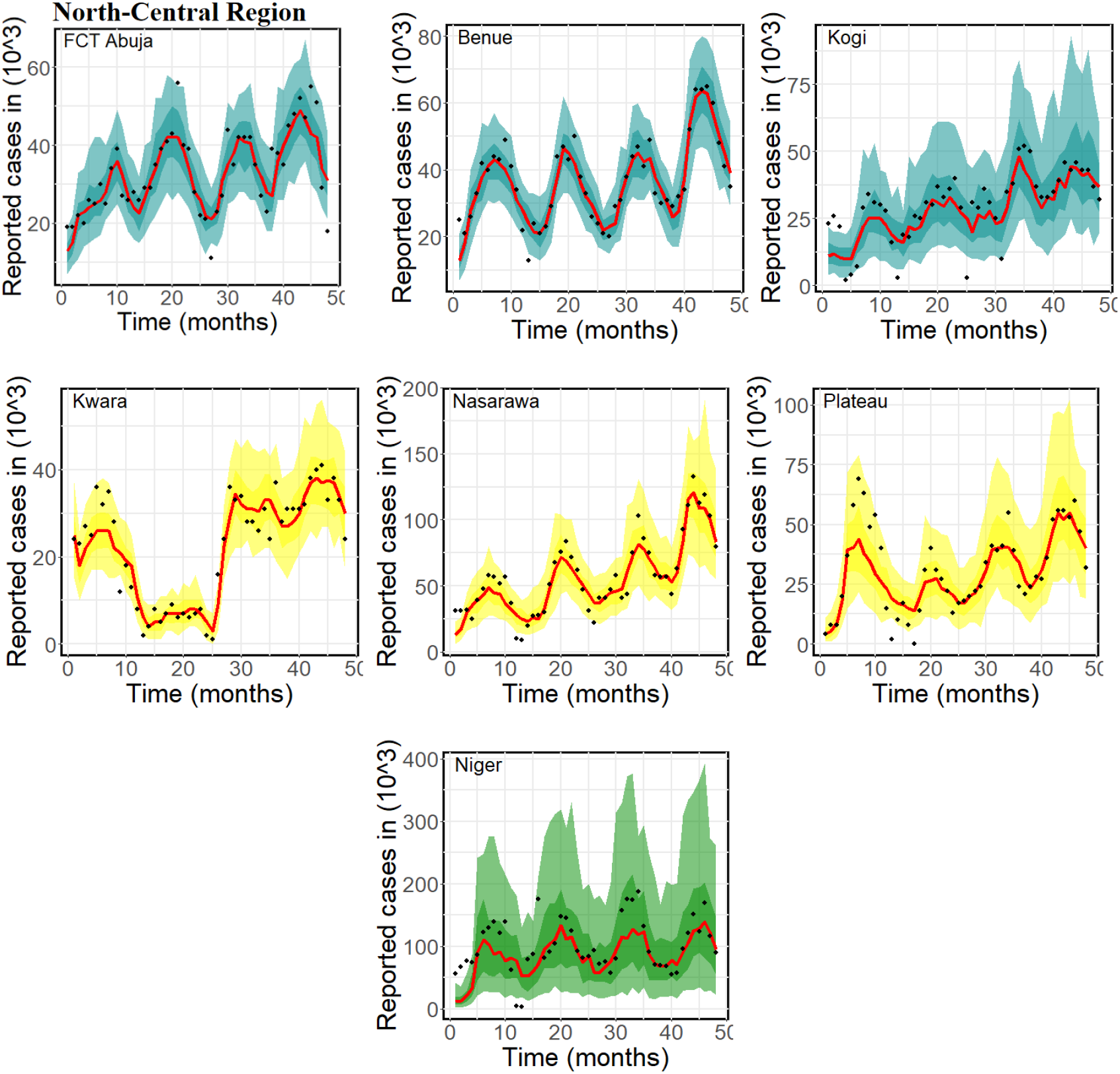
Model fit using the monthly reported malaria cases for the North Central Region consisting of Abuja, Benue, Kogi, Kwara, Nasarawa, Plateau and Niger states for the period of 4 years (January, 2014-December, 2017). The dotted line indicates the reported cases data while the red line indicates the model fit. The deeply coloured region represents 50% CrI while the lightly coloured region represents 95%CrI

In the top panel of Figure 7, we present Abuja with ℛ_*e*_=4.85 (95% CrI : 1.11-8.76), Benue with ℛ_*e*_=5.25 (95% CrI : 1.42-9.38) and Kogi with ℛ_*e*_=5.32 (95%CrI 0.45-10.98) from the North central region. In the bottom panel, we present the fit of Kwara, Nasarawa and Plateau states. The effective reproduction number of Kwara state was estimated to be 4.88 (95% CrI : 1.36-8.38), Nasarawa was 5.39 (95% CrI : 0.97-5.39) and that of Plateau was 4.50 (95% CrI : 0.67-9.17). Figure 8 shows the fit for the north-western region which comprises of Jigawa, Kaduna, Kano, Katsina, Kebbi, Zamfara and Sokoto states. The effective reproduction number of those states are estimated as 4.77 (95% CrI : 0.96-9.35), 5.25 (95% CrI : 1.01-10.49), 4.70 (95% CrI: 0.99-9.08), 5.03 (95% CrI : 1.14-9.66), 4.98 (95% CrI: 0.89-9.71), 4.63 (95% CrI: 1.01-8.48) and 4.93 (95% CrI: 0.64-9.31) respectively.

**Figure 8:**
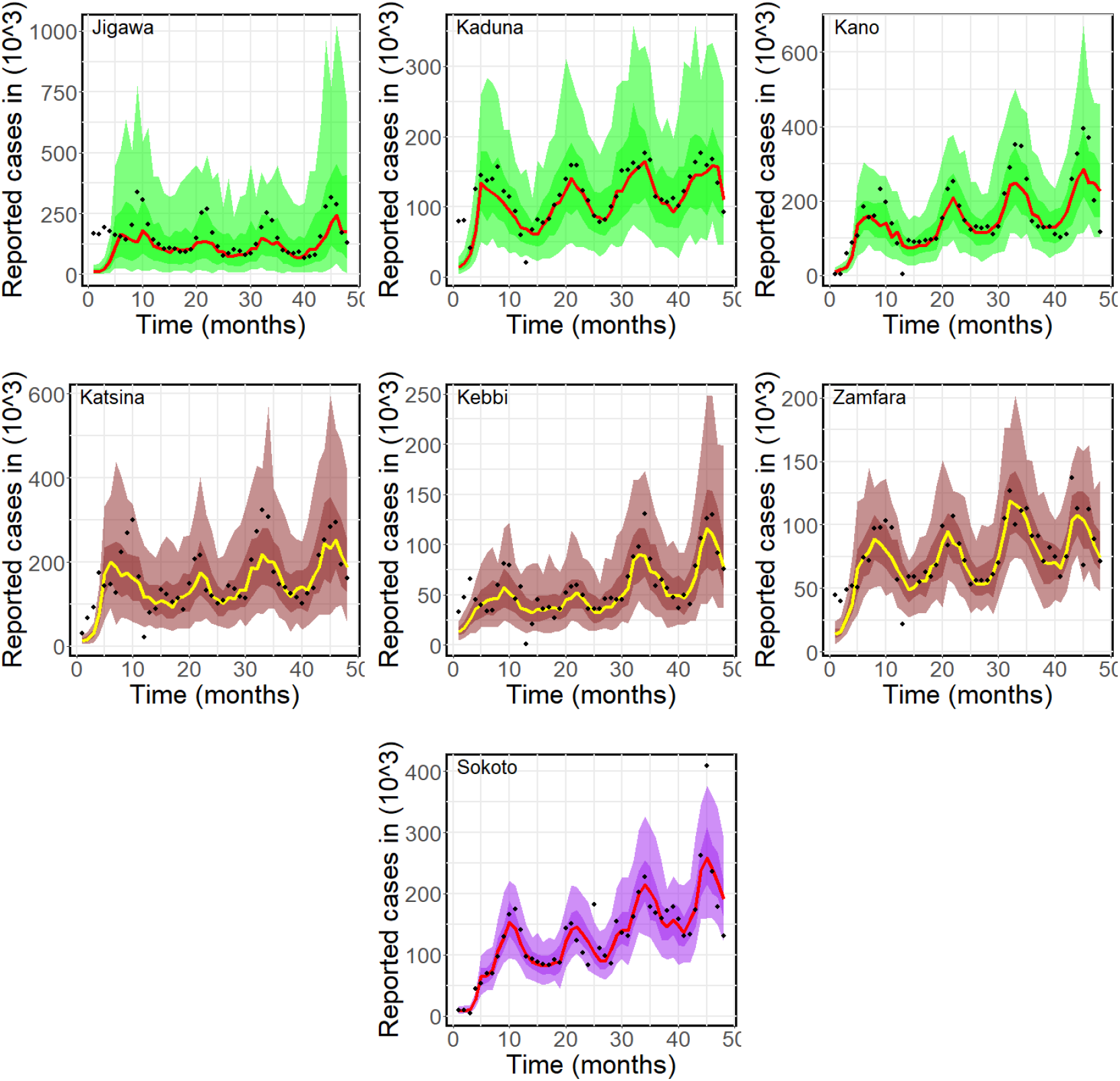
Model fit using the monthly reported malaria cases for the North Western Region consisting of Jigawa, Kaduna, Kano, Katsina, Kebbi, Zamfara and Sokoto states for the period of 4 years (January, 2014-December, 2017). The dotted line indicates the reported cases data while the red and yellow lines indicate the model fit. The deeply coloured region represents 50% CrI while the lightly coloured region represents 95%CrI

**Figure 9:**
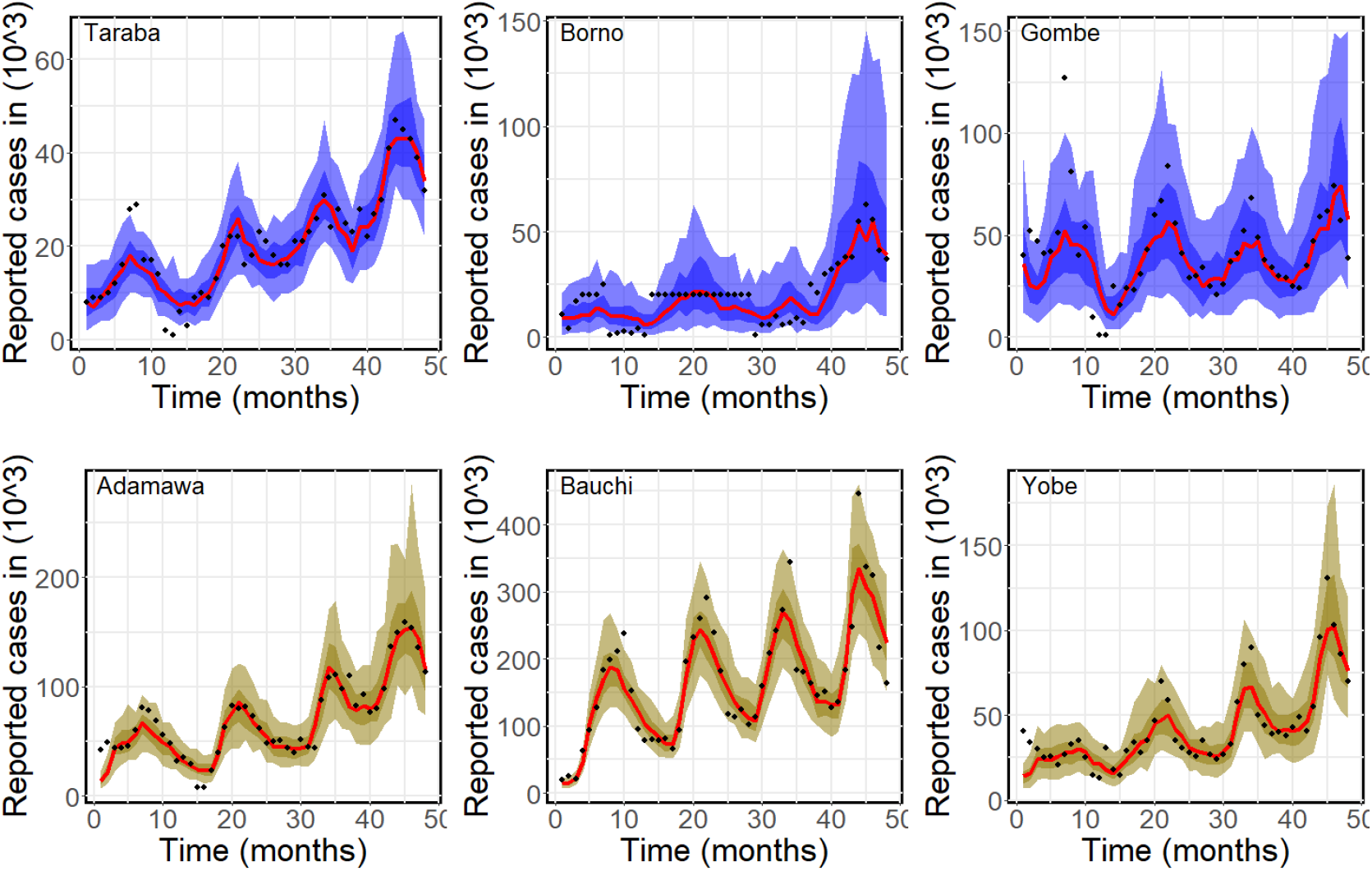
Model fit using the monthly reported malaria cases for the North-Eastern Region Consisting of Taraba, Borno, Gombe, Adamawa, Bauchi and Yobe states for the period of 4 years (January, 2014-December, 2017). The dotted line indicates the reported cases data while the red line indicates the model fit. The deeply coloured region represents 50% CrI while the lightly coloured region represents 95%CrI

In the north-central, the state with the highest effective reproduction number is Nasarawa 5.39 (95% CrI : 0.97-10.28) while the lowest in the region is Niger 4.25 (95% CrI : 0.89-8.09). In the north-west, the state with the highest effective reproduction number is Kaduna 5.25 (95% CrI : 1.01-10.41) while the lowest in the region is Zamfara 4.63 (95% CrI : 1.01-8.48). In the north-east, Adamawa has the highest reproduction number 5.92 (95% CrI : 1.60-10.59) and the lowest is Bauchi 3.72 (95% CrI : 1.11-7.08). Adamawa has the overall highest reproduction number while Bauchi is the lowest in the northern Nigeria. The effective reproduction number of the remaining states can be found in Table 3.

The estimated time-dependent contact rates of all the northern states are presented in Figures 10 -12. We present the mean estimated value of *C*(*t*) as 0.31 for Kaduna, Kano, Zamfara, Sokoto, Taraba and Yobe states. The value of 0.27 was estimated for Kogi, Benue and Adamawa states. Niger state has the highest *C*(*t*) 0.40 and the lowest was Gombe 0.26. The mean estimated contact rate for the other states can be found in Table 3 as well as their credible intervals. It was expected that Adamawa with the highest effective reproduction number should also have the highest contact rate instead of Niger but this was not the case. The reason for this is either the contact rate of Adamawa as estimated is far from the true contact rate due to the reason raised earlier about the reliability of the reported cases data in Adamawa or it is because the mosquito abundance in Niger is more than that of Adamawa state.

**Figure 10:**
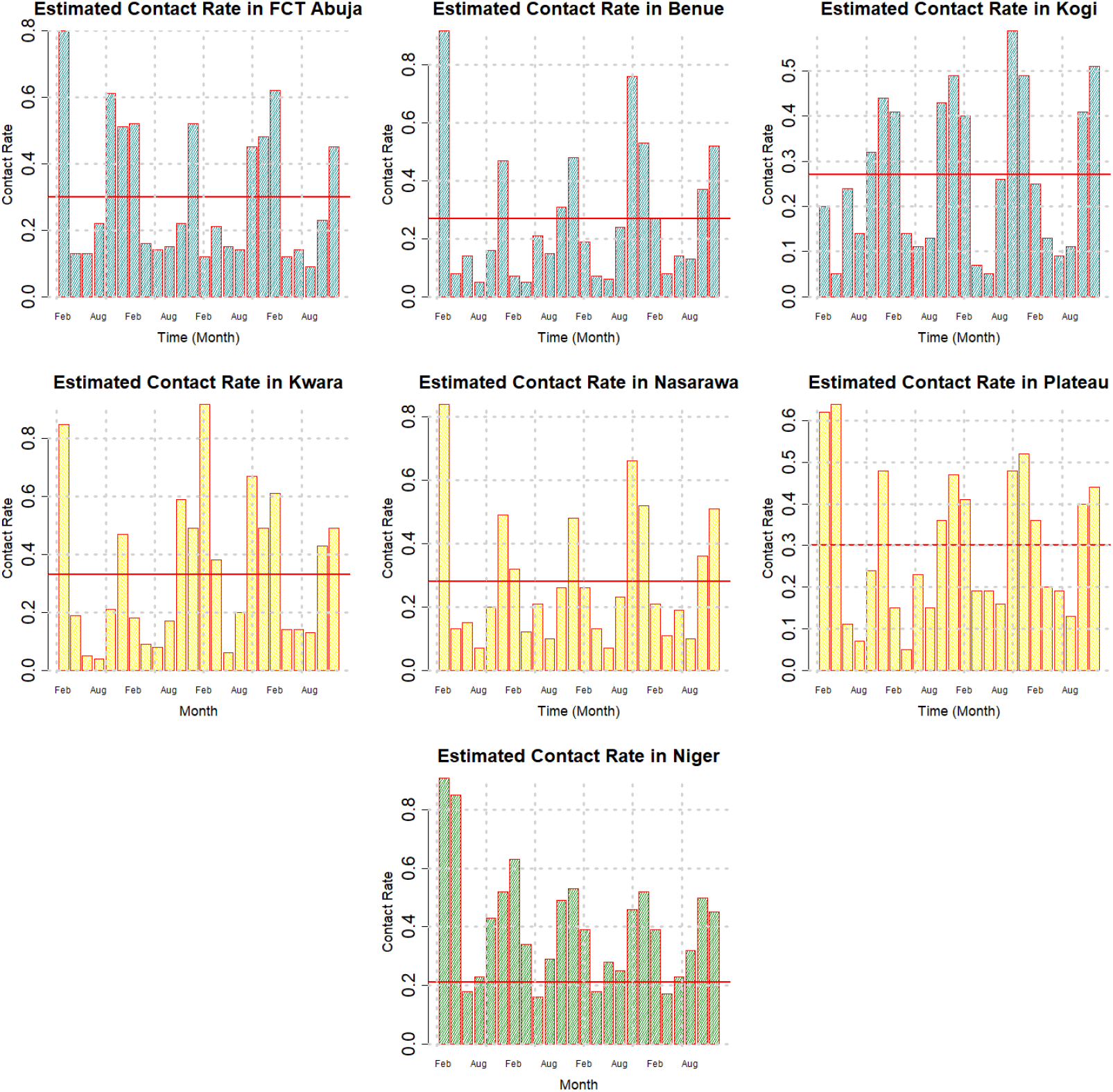
Plots of the estimated contact rates for the North Central Region consisting of Abuja, Benue, Kogi, Kwara, Nasarawa, Plateau and Niger states. The red line shows the average. Each data point corresponds to two months starting from January 2014.

**Figure 11:**
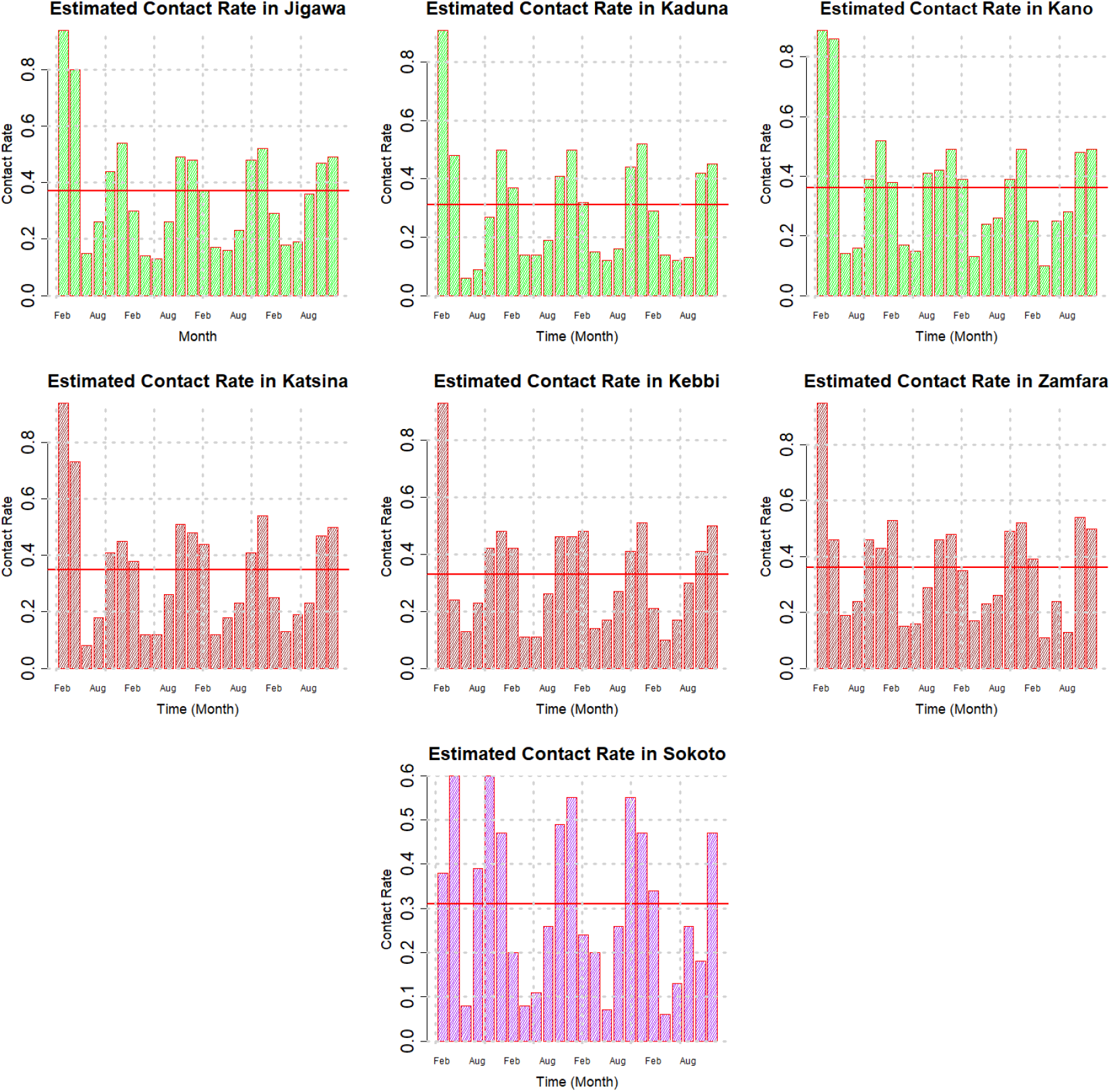
Plots of the estimated contact rates for the North Western Region consisting of Jigawa, Kaduna, Kano, Katsina, Kebbi, Zamfara and Sokoto states. The red line shows the average. Each data point corresponds to two months starting from January 2014.

### 5.3 Estimated Time-dependent Contact Rate

Figures 13 -15 were plotted using the estimated contact rates in Figures 10 -12 respectively. In order to fit the model to the reported cases data such that the model will be able to capture the waves and pattern of the data, we assumed that the contact rate between human and mosquito changes over time. This consequently leads to changes in the biting rate, transmission rate and the reproduction number. The fitting process reveals that there is bimonthly changes in the contact rate since the reported cases data is monthly. So doing, we estimated 24 contact rates for each states since there exists only 48 data points between January 2014 and December 2017. The estimated time-dependent contact rates and the reproduction numbers are presented in Figures 10 -12 and 13 -15 respectively. The red line shows the mean value.

**Figure 12:**
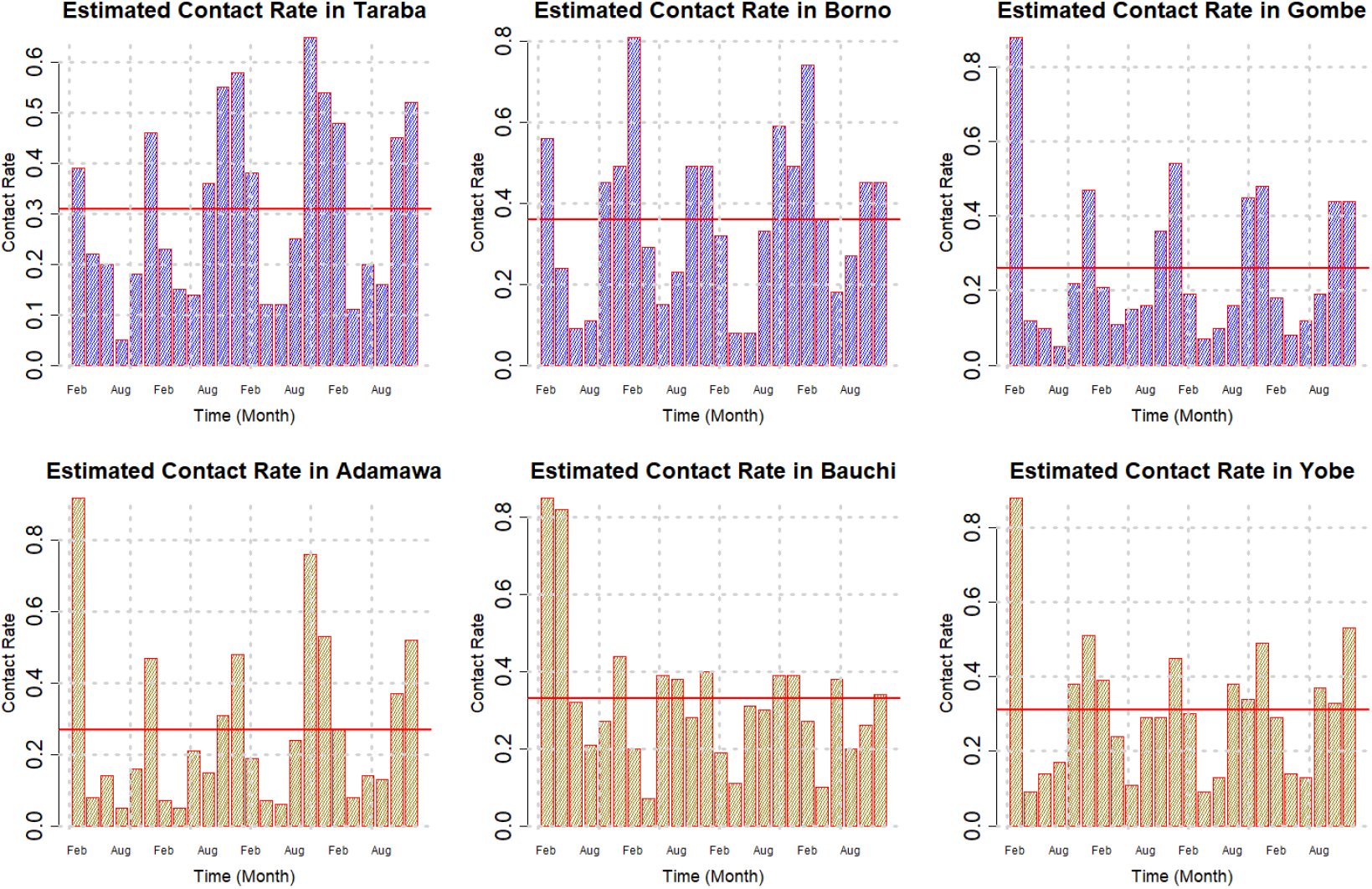
Plots of the estimated contact rates for the North-Eastern Region Consisting of Taraba, Borno, Gombe, Adamawa, Bauchi and Yobe states. The red line shows the average. Each data point corresponds to two months starting from January 2014.

**Figure 13:**
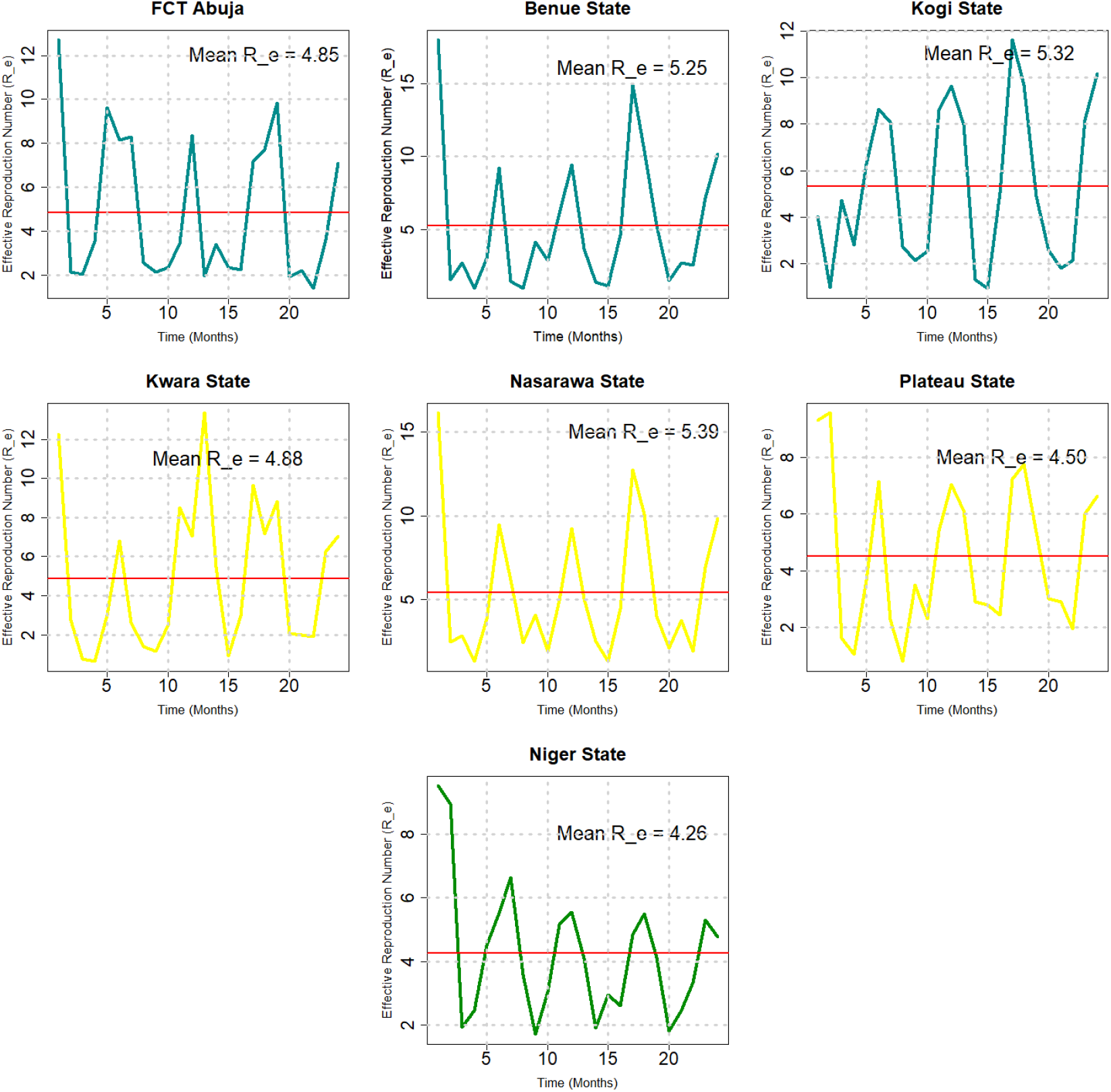
Plots of the estimated reproduction numbers for the North Central Region consisting of Abuja, Benue, Kogi, Kwara, Nasarawa, Plateau and Niger states. The red line shows the mean. Each data point corresponds to two months starting from January 2014.

### 5.4 Estimated effective reproduction number

It can be observed in Figures 13 -15, in the majority of the northern states that the reproduction number is very high between November and December (up to February in some states). In fact the peak of the reproduction number occurs in January in the majority of the states (see Kwara 13.5, Nasarawa 17.5, Benue 18, Plateau 9.5, Gombe 18.5 among others).

**Figure 14:**
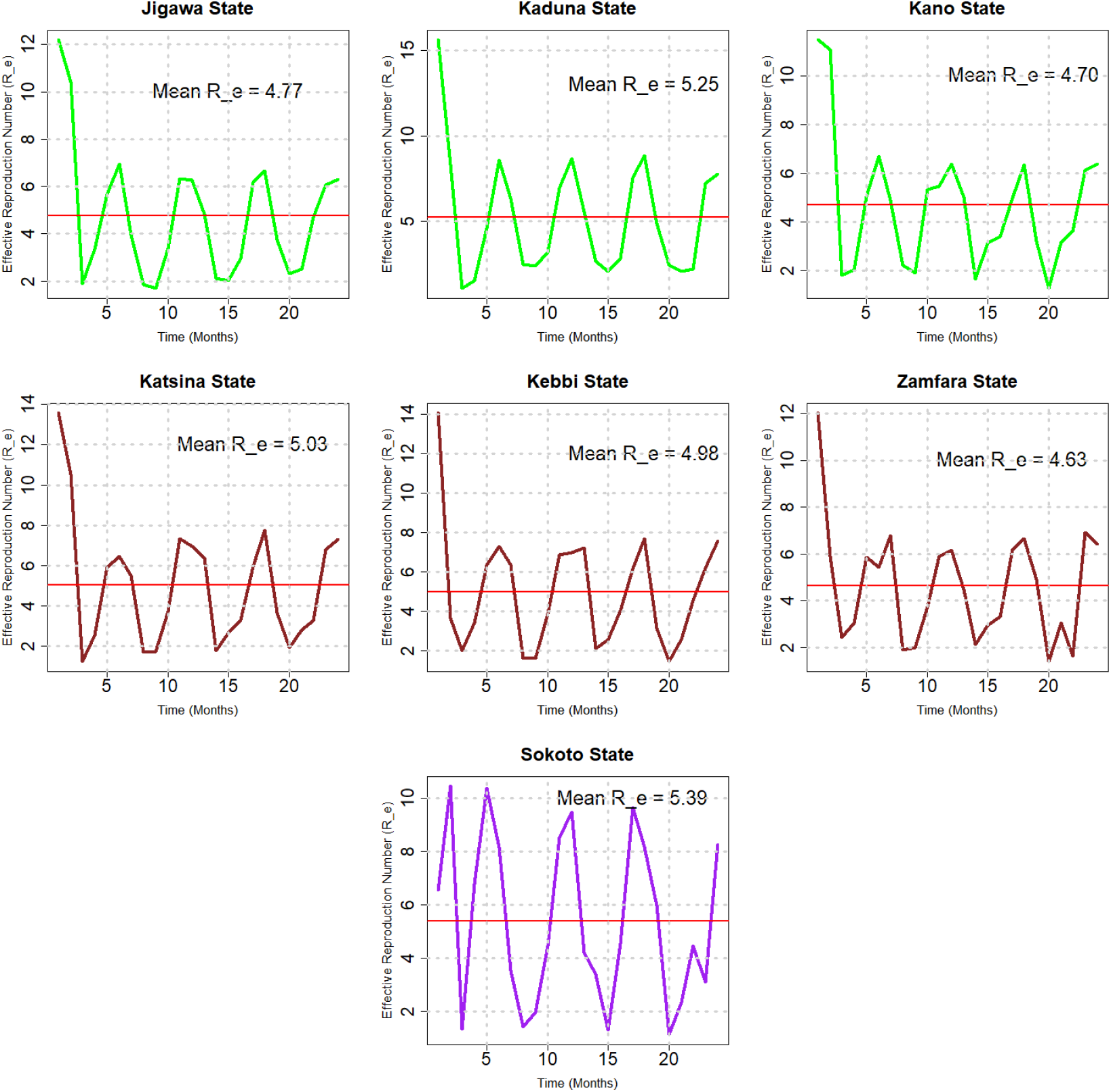
Plots of the estimated reproduction numbers for the North Western Region consisting of Jigawa, Kaduna, Kano, Katsina, Kebbi, Zamfara and Sokoto states. The red line shows the mean. Each data point corresponds to two months starting from January 2014.

**Figure 15:**
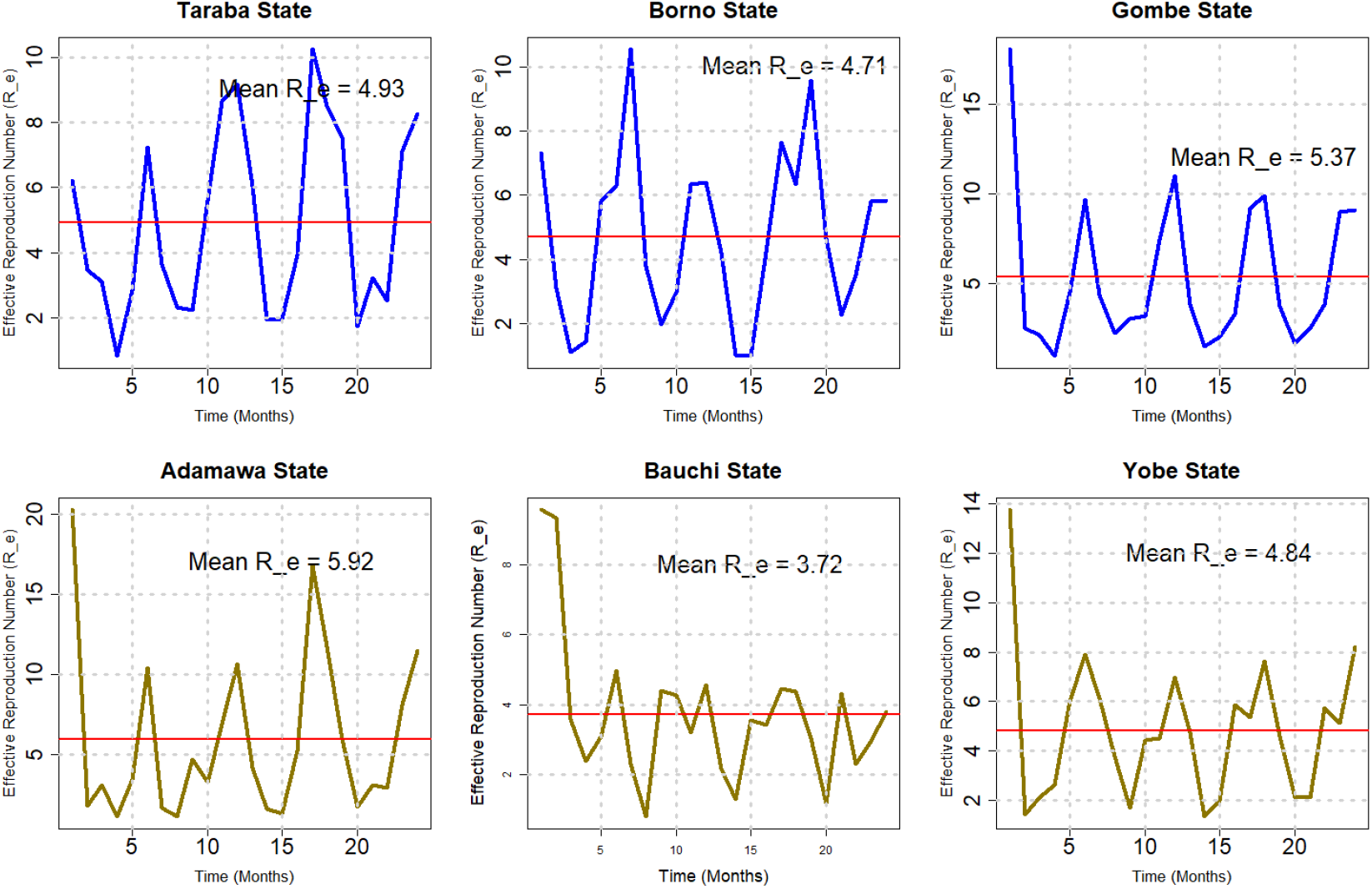
Plots of the estimated reproduction numbers for the North-Eastern Region Consisting of Taraba, Borno, Gombe, Adamawa, Bauchi and Yobe states. The red line shows the mean. Each data point corresponds to two months starting from January 2014.

The lowest value of the reproduction number occurs averagely between March and June in the majority of the states such as Bauchi 0.39, Borno 0.2, Gombe 0.36, Sokoto 0.4 among others.

One interesting result that can be deduced here is that the abundance of mosquito and malaria in Nigeria is season-dependent. The reproduction number is very high between March/April and October/November which corresponds to rainy season in Nigeria while the lowest reproduction number being estimated falls between December and February which corresponds to the dry season (though, the reproduction number is also high around February in some states in 2014).

In Figure 16, It can be seen vividly that the contact rate is not directly proportional to the reproduction number. For instance, the states with the highest reproduction number are Adamawa, Nasarawa, Gombe, Kogi, Benue, Kaduna etc while the states with the highest contact rate are Niger, Jigawa, Borno, katsina etc. This means that high contact rate does not always guarantee high reproduction number. This is because there are other factors such as recovery rate, number of bites, probability of infectious bites that are heavily dependent on the reproduction number.

**Figure 16:**
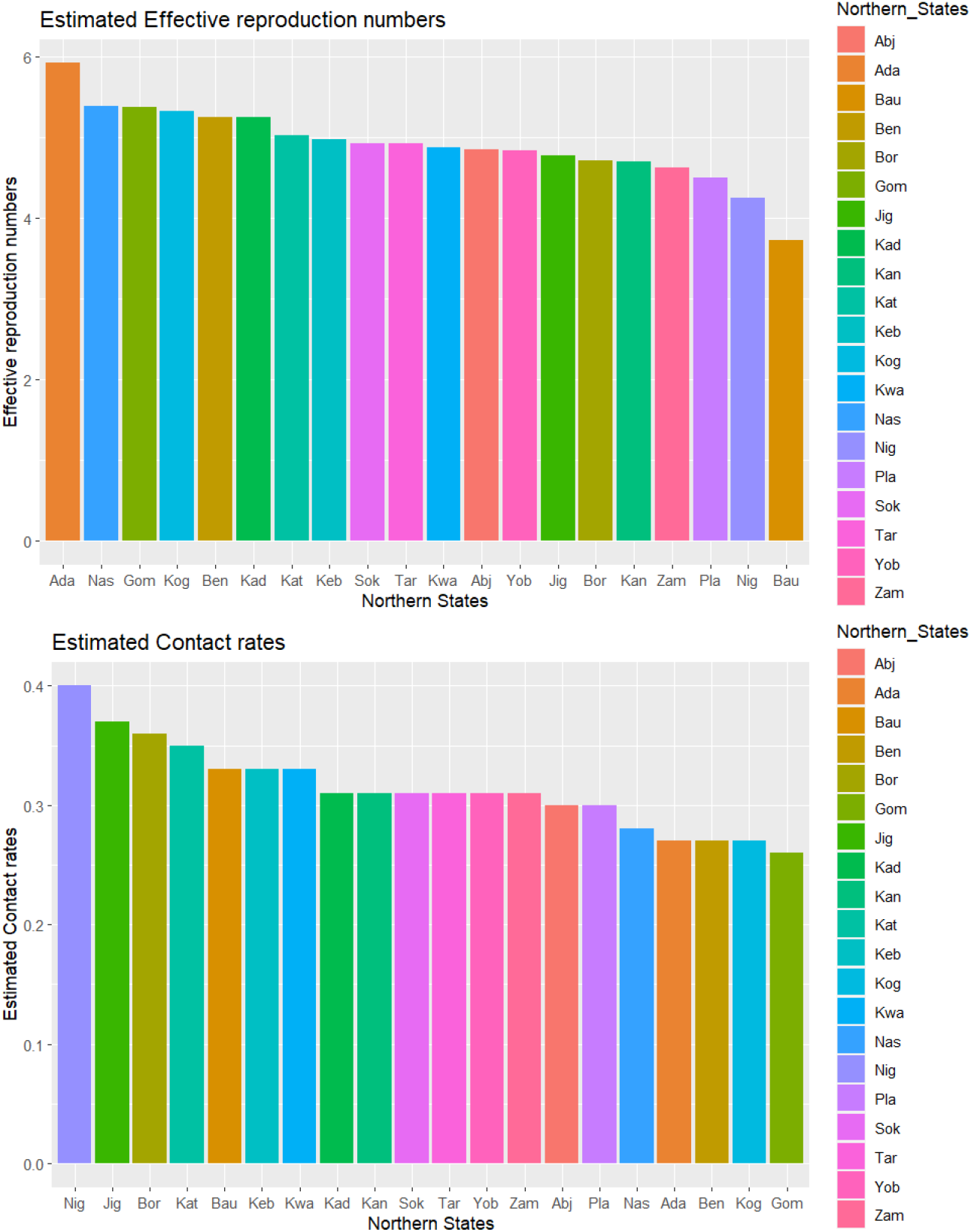
Plots of the estimated effective reproduction numbers and contact rates for all northern states.

## Conclusion

Often infected by a particular species of mosquito that feeds on humans, malaria is a dangerous and occasionally fatal disease brought on by a parasite. Despite all forms of interventions such as LLIN, SMC, IRS being implemented in Nigeria and worldwide, malaria still remains endemic spreading across the country. In this study, we used a mathematical modelling technique to estimate the effective reproduction number (which is one of the most important epidemiological threshold) of malaria in all the northern states. So doing, we estimated 24 time-dependent contact rates for all the 20 northern states. We used the reported malaria cases data obtained from the national malaria elimination programe NMEP and demographic data. A Bayesian statistical framework with rstan written in C++ was used for the model validation, parameter estimation and fitting.

The main findings of this research are summarized as follows:

1. the proposed mathematical model (incorporating seasonality, super-infection and time-dependent contact rate) was able to predict the trend, wave and seasonal variation of the reported malaria cases data. With an exception of the initial take-off of Jigawa, Gombe, Niger, Zamfara, Yobe and Adamawa states, this anomaly can be attributed to over-reporting of the malaria cases data during the initial spread,
2. the posterior mean obtained from the Bayesian inference shows that in the north-central, the state with the highest effective reproduction number is Nasarawa 5.39 (95% CrI : 0.97-10.28) while the lowest in the region is Niger 4.25 (95% CrI : 0.89-8.09). In the north-west, the state with the highest effective reproduction number is Kaduna 5.25 (95% CrI : 1.01-10.41) while the lowest in the region is Zamfara 4.63 (95% CrI : 1.01-8.48). In the north-east, Adamawa has the highest reproduction number 5.92 (95% CrI : 1.60-10.59) and the lowest is Bauchi 3.72 (95% CrI : 1.11-7.08). Adamawa has the overall highest reproduction number while Bauchi is the lowest in the northern Nigeria.
3. Adamawa state has the highest reproduction number while Niger has the highest contact rate. The reason for this is probably because the contact rate of Adamawa as estimated is far from the true contact rate due to the reliability of the reported cases data in Adamawa or it is because the mosquito abundance in Niger is more than that of Adamawa state,
4. the Contact rate is not directly proportional to the reproduction number,
5. according to the available data, it was derived during the analysis that over 60% of the malaria cases in the northern Nigeria are attributed to the asymptomatic individuals,
6. the effective reproduction number is high during the rainy season (March/April-October/November) while it is low during the dry season (December-March). It is also high in February in some states in 2014 because those states experience minimal rainfall in February.

The findings of this study will contribute to the development of mathematical models that are used to examine the dynamics of malaria in the chosen states. Additionally, public health professionals and government representatives can use the time-dependent contact rate and the effective reproduction number estimates to properly plan and outline tactics that will aid in the eradication of the disease.

This study is not without limitations. One of the major challenges is availability of quality data in Nigeria. The available data for this study was only between January 2014 and December 2017. The model did not consider the age heterogeneity ie. the estimated quantities are not taken down to the age-structure level where proper understanding of the age stratification can be achieved. The geo-spatial analysis of the selected states are not taken into consideration ie. the selected states are not considered at the local-government and district level. The proposed model only considers seasonal patterns in the contact rate but does not consider it in either the birth or death rate of the mosquitoes as these rates are also seasonal. Our future direction is to examine one or more of these limitations in order to advance the scope of this research.

## Data Availability

All data produced in the present study are available upon reasonable request to the authors and NMEP

## Acknowledgement

We acknowledge funding from the Bill and Melinda Gates Foundation through WAMCAD and ICAMMDA. We also acknowledge the Malaria Consortium and NMEP for giving us the data.

## Data Availability

All the cases data used in this research are available with the national malaria elimination programe [34].

## A Appendix

**Figure S1:**
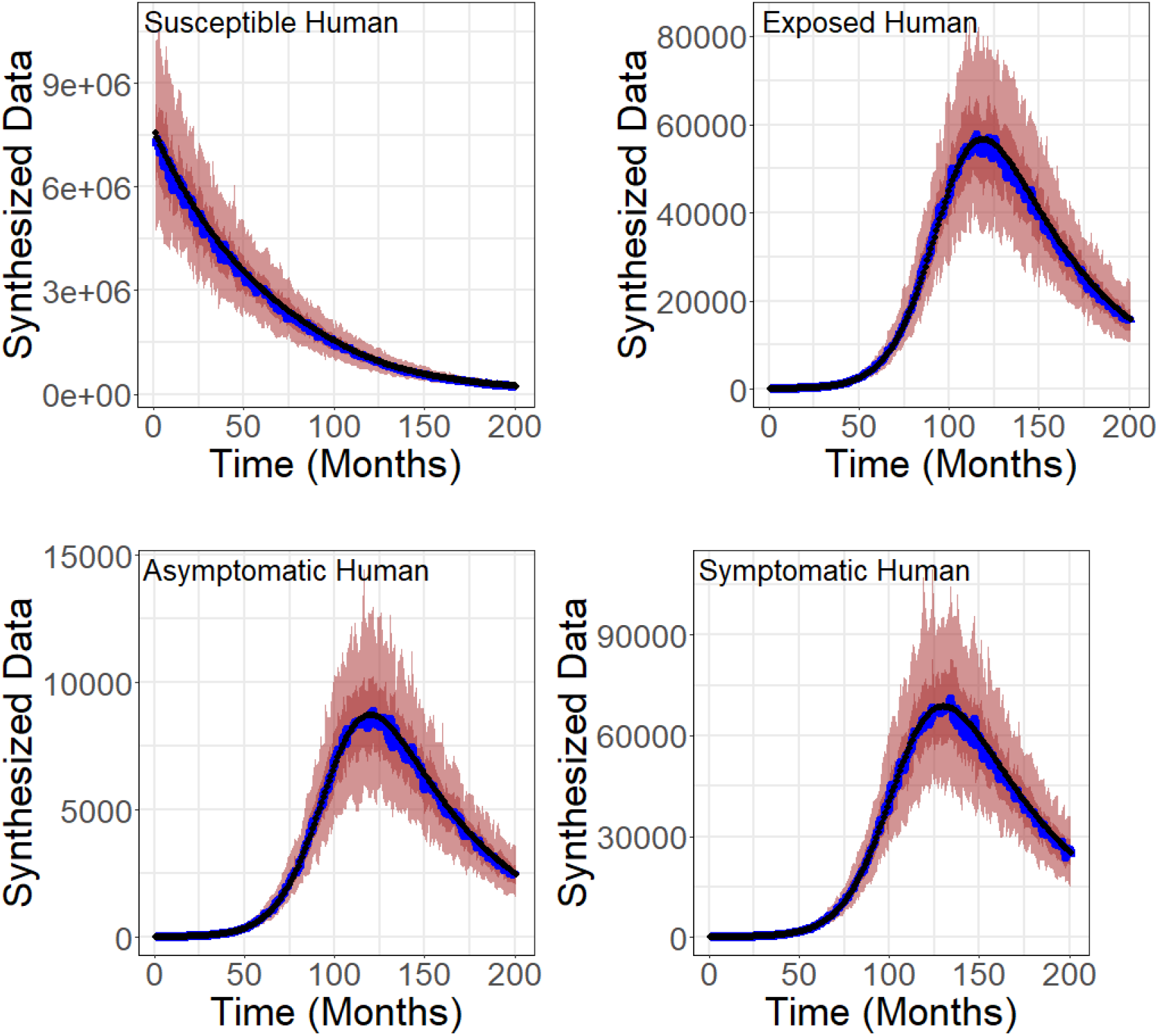
Model fit for the model equation. (2.3) - (2.6) using synthesized data. The black line indicates the synthesized data while the blue line indicates the model fit. The deeply coloured part represents 50% CrI while the lightly coloured part represents 95%CrI.

**Figure S2:**
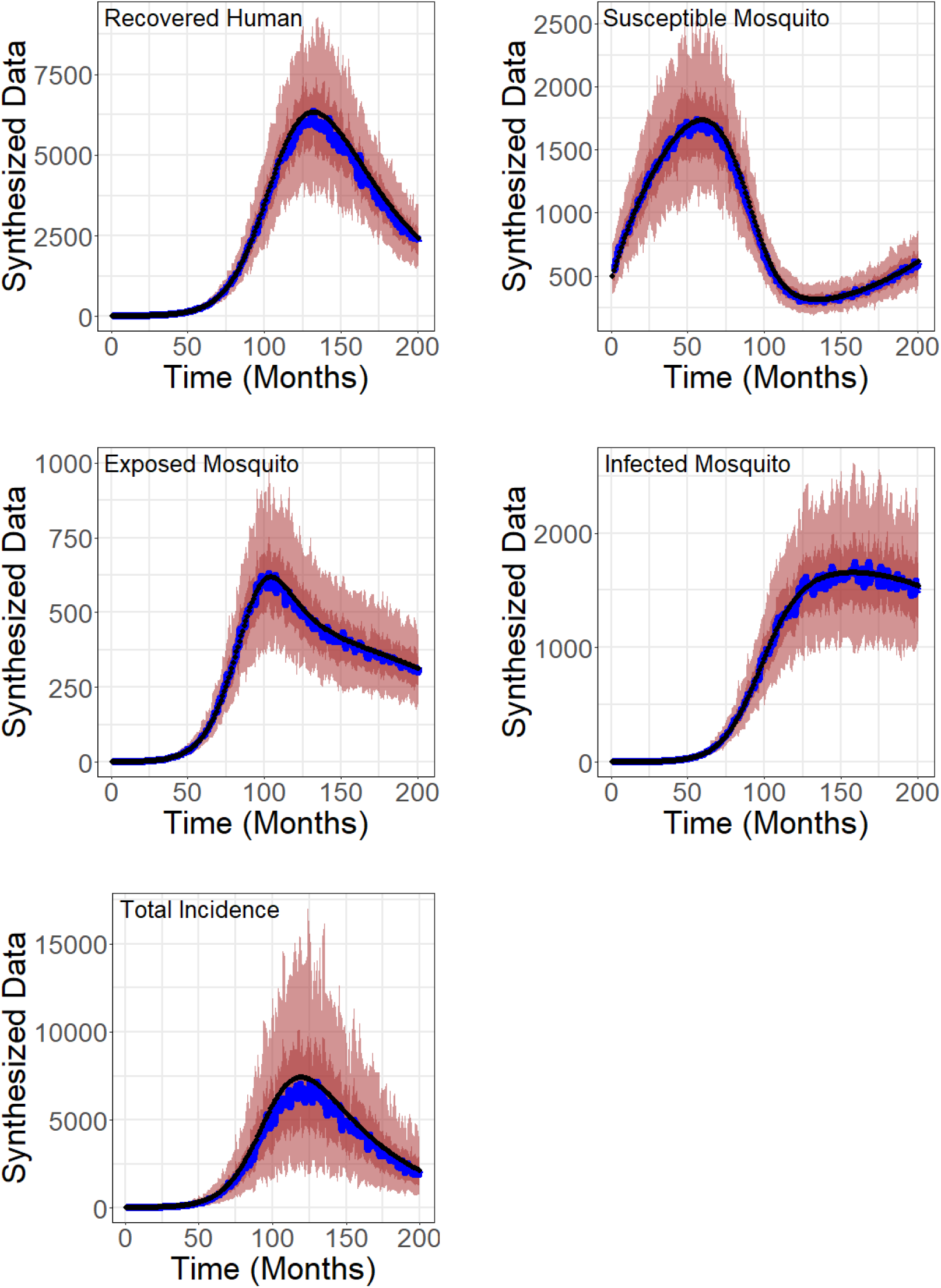
Model fit for the model equation. (2.3) - (2.6) using synthesized data. The black line indicates the synthesized data Sw-3hile the blue line indicates the model fit. The deeply coloured part represents 50% CrI while the lightly coloured part represents 95%CrI.

